# “Actionable” Risk for Preterm Birth: Patterns and Prediction in California Singleton Births 2016-2020

**DOI:** 10.1101/2025.11.14.25340227

**Authors:** Laura L. Jelliffe-Pawlowski, HOPE Lab, Rebecca J. Baer, Scott Oltman, Safyer McKenzie-Sampson, Deborah Adeyemi, Ashley Becker, Kacie C.A. Blackman, Bridgette Blebu, Justin S. Brandt, Elena Flowers, Dana R. Gossett, Emily C. Hanselman, Sasha Hernandez, Liang Liang, Audrey Lyndon, Allison M. Momany, Elizabeth E. Rogers, Kelli K. Ryckman, Louie M. Swander, Karen M. Tabb, Kelly D. Taylor, Sophia L. Wiggins, Akila Subramaniam

**Affiliations:** Rory Meyers College of Nursing, New York University, New York, New York; Department of Obstetrics & Gynecology, Grossman School of Medicine, New York University, New York, New York; Department of Obstetrics & Gynecology, New York University Langone Health, New York, New York; Department of Epidemiology and Biostatistics, University of California San Francisco, San Francisco, California; Department of Global Health Sciences, University of California San Francisco, San Francisco, California; The California Preterm Birth Initiative, University of California San Francisco, San Francisco, California; Healthy Outcomes of Pregnancy for Everyone (HOPE) Research Consortium, University of California San Francisco, San Francisco, CA; New York University, New York, New York; UCSD Study of Outcomes in Mothers and Infants (SOMI), La Jolla, CA; Department of Obstetrics, Gynecology and Reproductive Sciences, University of California San Francisco, San Francisco, California; Department of Pediatrics, University of California, San Francisco, San Francisco, California; Department of Pediatrics, University of California San Diego, La Jolla, California; Department of Health Behavior and Health Equity, School of Public Health, University of Michigan, Ann Arbor, Michigan; Department of Family Community Medicine, University of California, San Francisco, San Francisco, California; Department of Health Sciences, California State University Northridge, Northridge, California; Department of Obstetrics and Gynecology, The Lundquist Institute at Harbor-UCLA Medical Center, Torrance, CA; School of Nursing, University of California San Francisco, San Francisco, California; Department of Obstetrics and Gynecology, Medical College of Wisconsin, Milwaukee, Wisconsin; Stead Family Department of Pediatrics, University of Iowa, Iowa City, Iowa; Department of Epidemiology and Biostatistics, School of Public Health-Bloomington, Indiana University, Bloomington, Indiana; School of Social Work, University of Illinois Urbana-Champaign, Urbana, Illinois; Department of Medicine, University of California, San Francisco, San Francisco, California; Department of Obstetrics and Gynecology, University of Alabama Birmingham, Birmingham, Alabama

## Abstract

**Background:** Preterm birth (PTB, <37 weeks of gestation) is the leading cause of child mortality in the United States (U.S.) and worldwide, and has substantial short– and long-term health consequences for mothers and infants. Each year, >350,000 infants in the U.S. are born preterm, and rates continue to rise in parallel with maternal risk factors such as hypertension, diabetes, anemia, asthma, and mental health conditions. Evidence-based interventions exist for many of these conditions and are associated with improved pregnancy outcomes, including low-dose aspirin for preeclampsia prevention in individuals with chronic hypertension or pregestational diabetes, inhalers for asthma, iron for anemia, and therapy or medication for mental health disorders, but fewer than half of eligible individuals receive them, reflecting persistent gaps in use. To address this, we developed the PTB Actionable Risk Index (PTB-ARIx), which leverages factors with known evidence-based interventions to identify individuals who are pregnant and are at increased risk for PTB. This study evaluates performance of the PTB-ARIx throughout pregnancy in terms of risk determination and characterization of actionable risk factors, including their combined contributions to PTB.

**Methods and Findings:** A retrospective cohort study was conducted using linked data for 1.9 million singleton live births in California in 2016-2020, divided into training and testing sets. Poisson regression estimated associations between 18 candidate risk factors for PTB with evidence-based interventions spanning clinical, behavioral, and social risks, including preeclampsia risk composites (≥1 high-risk or ≥2 moderate-risk factors based on U.S. Preventive Services Task Force (USPSTF) criteria), maternal conditions (e.g., gestational hypertension, asthma), substance use, and social adversity. Beta coefficients were combined to construct the PTB-ARIx, evaluated by per-unit associations with PTB and by area under the receiver operating characteristic curve (AUC) overall, by early (<32 weeks), late (32-36 weeks), spontaneous, and medically indicated PTB, and by PTB co-occurring with preeclampsia.

All risk factors were found to be associated with increased PTB risk. Having ≥1 high-risk or ≥2 moderate-risk factors for preeclampsia (based on composites) was most strongly related to PTB (relative risk (RR) 6.73, 95% confidence interval (CI) 6.57, 6.89). Each unit increase in PTB-ARIx was associated with >60% higher PTB risk (RRs 1.66–1.72) across training and testing samples, with consistent findings across PTB and race/ethnicity–insurance subgroups. Model performance was modest for late PTB (AUC ≈0.63), stronger for early PTB (0.69–0.72), and especially high for early PTB with preeclampsia (AUCs up to 0.97). Over 70% of individuals with PTB-ARIx scores ≥3.00 experienced PTB or another adverse outcome such as low birth weight (<2500 grams).

**Conclusions:** The PTB-ARIx is a well-performing metric for identifying individuals at increased risk for PTB and other adverse pregnancy outcomes. By centering on modifiable risks, the PTB-ARIx combines risk identification with opportunities for intervention. Demonstrating strong performance across subgroups, including for early PTB and PTB with preeclampsia, the PTB-ARIx provides a potential pathway to improve patient–provider communication and uptake of equitable, evidence-based care. Further validation, including integration with treatment data, is needed to confirm its potential to reduce PTB risk and rates.

**Author Summary:** *Why was this study done?:*

- Preterm birth (PTB), or delivery before 37 weeks of pregnancy, is a leading cause of newborn illness and death worldwide, and rates are rising in parallel with increases in known risk factors like hypertension, diabetes, asthma, anemia, and mental health conditions.
- Effective, evidence-based treatments for known PTB risk factors are underutilized.
- Many existing tools predict PTB using statistical thresholds but do not highlight risk factors with proven treatment(s) or intervention(s) during pregnancy.
- There is a need for approaches that both predict PTB and link directly to actions that can reduce risk.

*What did the researchers do and find?:*

- We used health data from more than 1.9 million births in California to develop the Preterm Birth Actionable Risk Index (PTB-ARIx).
- The PTB-ARIx included 18 risk factors grouped into: (1) composite preeclampsia risk groups (≥1 high-risk factor or ≥2 moderate-risk factors, as defined by U.S. Preventive Services Task Force guidelines), (2) maternal medical conditions (such as prior PTB, gestational diabetes, asthma, and anemia), (3) infections and reproductive health (such as sexually transmitted or urinary tract infections), (4) behavioral risks (such as smoking and substance use), and (5) social and care-related risks (such as food insecurity, and housing instability).
- The PTB-ARIx showed consistent performance in predicting different types of PTB, including early PTB and PTB with preeclampsia, with similar performance across race/ethnicity and insurance groups.
- We also found that the number of prenatal visits partly explained some of the relationship between risk scores and outcomes, suggesting that regular care may play a role in mitigating PTB risk.

*What do these findings mean?:*

- The PTB-ARIx provides a new way to predict PTB that highlights risk factors where preventive treatments or interventions, such as aspirin for individuals at increased risk of preeclampsia, can be applied during pregnancy.
- This model may help providers and patients work together to better identify, understand, and reduce risk, supporting more equitable care across diverse populations.
- Further research is needed to test the tool in other settings, study how treatments affect risk, and evaluate whether a patient-facing version can improve uptake of interventions and pregnancy outcomes.

## Introduction

Preterm birth (PTB), birth before 37 weeks of gestation, is the leading cause of child mortality in the United States (U.S.) and globally [1,2]. Each year, there are more 350,000 PTBs in the U.S. [3], driving health care costs that exceed $25 billion [4]. Despite decades of research investment and clinical focus [5], PTB rates have increased. Between 2016 and 2023, the PTB rate among singleton births rose from 8.0% to 8.7% in the U.S. [3].

PTB carries profound health consequences for both pregnant individuals and infants. Individuals who deliver preterm face higher risks for postpartum infection, preeclampsia, and severe maternal morbidity (SMM) [6–9], as well as elevated long-term risks for cardiovascular and renal disease, cancer, and all-cause mortality [10–17]. Infants born preterm are at increased risk of neonatal and infant death [2,18,19] and, as they age, are more likely to develop chronic conditions such as asthma, diabetes, hypertension, and stroke [20–25]. They also may experience elevated risks for neurodevelopmental delays and mental health disorders, including anxiety and depression, that may develop during childhood and adolescence and persist throughout adulthood [26–32].

The burden of PTB is unequally distributed and closely tied to social determinants of health (SDoH), including race and ethnicity, income, and other factors related to structural determinants due to policies and income inequities.

Disparities in outcomes among individuals who give birth preterm and those who give birth at term, and their infants, have continued to widen, particularly at the intersection of geography, race/ethnicity, and income [3,33,34–37].

Inequities in the burden of PTB extend beyond delivery. Low-income Black/African American individuals who deliver preterm are more likely to experience SMM [36], while their infants face higher risks for bronchopulmonary dysplasia (BPD), retinopathy of prematurity (ROP), and mortality, even when born at similar gestational ages as infants from other groups [37].

### Risk Factors, Evidence-Based Interventions, and Underutilization

Growing evidence links rising rates in key risk factors to both the overall increase in PTB and widening inequities. In a study of more than 5.1 million singleton births in California from 2011–2022, PTB rates were found to have climbed from 6.8% to 7.5%, a 10.6% relative increase [34]. This increase mirrored sharp upticks in chronic and gestational hypertension, preexisting and gestational diabetes, prior PTB, obesity, asthma, anemia, autoimmune diseases, infections complicating pregnancy, mental health conditions, substance use, and housing insecurity across race, ethnicity, and insurance status [34]. The study also found that some of the largest increases in the prevalence of risk factors were concentrated in populations already experiencing PTB disparities. For instance, from 2016–2022, gestational hypertension rose 51.8% (from 8.5% to 12.9%) in non-Hispanic (NH) Black/African American individuals with MediCal (California’s Medicaid) and 103.9% (from 7.7% to 15.7%) in those without MediCal [34]. National CDC data show similar trends: between 2016–2023, gestational hypertension rates rose 60.5% (from 7.2% to 11.5%) in NH Black/African American individuals with Medicaid and 62.3% (7.1% to 11.5%) in those without Medicaid [3,35].

While several evidence-based interventions targeting risk factors closely linked to PTB have been shown to reduce the risk of PTB and improve birth outcomes, uptake remains suboptimal across multiple risk factors and groups (Supplementary Table 1) [38–165]. Recommended interventions that may be prescribed by providers include, for example, the use of low-dose aspirin after 12 weeks’ gestation for individuals with preexisting diabetes or chronic hypertension as preeclampsia prophylaxis in pregnancy [38]; pharmacologic management of asthma [72]; and therapeutic and/or pharmacologic treatment for mental health conditions during pregnancy [125–127]. Documented gaps in utilization are large. Multiple independent studies have highlighted low uptake of evidence-based interventions among pregnant individuals with specific risk factors. In one study, only 57% of those at high risk for preeclampsia (e.g., preexisting diabetes, chronic hypertension, or prior preeclampsia) received low-dose aspirin [40]. In a separate study of pregnant individuals with asthma, just 32% reported use of inhalers or other medications [75]. Another study found that among those with a diagnosed mental health condition, as few as one in three (33%) received treatment [135].

Marked disparities have also been observed in evidence-based intervention use across race/ethnicity and socioeconomic groups. In one study, the use of continuous glucose monitoring (CGM) among individuals with type 1 diabetes was found to be 31.1% in those covered by private insurance, compared to 9.6% among those with public insurance [44]. In another study, treatment rates for mental health conditions during pregnancy ranged from 19.1% among NH Black/African American individuals to 40.7% among NH White individuals [135].

### Creating an Actionable Risk Index for Preterm Birth: Closing “Know-Do” Gaps

Rising rates of PTB [1,3], growing prevalence of associated risk factors [34,35], and the persistent underutilization of evidence-based interventions (sometimes referred to as “know-do gaps” [166]) underscore the urgent need for more effective strategies to identify and respond to risk. Critically, the consistently low uptake of interventions known to improve maternal health and reduce the risk of PTB and related complications, such as preeclampsia, represents a missed opportunity for both prevention and protection, particularly in groups experiencing health inequities. These gaps offer clear inroads for strategies that may hold particular promise for improving the uptake of evidence-based interventions and ultimately reducing PTB rates in those at the highest risk.

One such opportunity lies in the development and implementation of an actionable PTB risk index for patients and providers, an approach that considers clinical, behavioral, and social risks with related evidence-based interventions. Such an index could be used to assist in discussions of risk and related interventions between pregnant people and their providers. Shifting toward a risk identification framework rooted in actionability would mark a departure from current prenatal care practices, where formal PTB risk scoring is rarely used. Some prenatal care providers have expressed reluctance to discuss elevated PTB risk without having clear, evidence-based interventions to offer, citing concerns that such conversations could increase patient anxiety, foster blame, erode trust, or reinforce a culture of risk [167,168]. In contrast, patients have reported valuing transparency about risk for PTB and other adverse outcomes, even in the absence of definitive treatments, to feel informed, prepared, and more actively engaged in their care [167,169–172]. This disconnect may help explain why existing PTB prediction tools [173–178], particularly those that emphasize nonmodifiable factors or rely on inputs with limited translational value, have seen limited uptake in clinical settings.

Focusing on actionable risks that can be addressed during pregnancy may help bridge this communication divide. Such a focus has the potential to facilitate earlier and more targeted uptake of evidence-based interventions, improve communication between patients and providers, and ultimately reduce the risk of PTB and other closely linked outcomes (e.g., preeclampsia, low birth weight (LBW; <2500 grams)). This approach also aligns with recent calls from the American College of Obstetricians and Gynecologists (ACOG) and the Society for Maternal-Fetal Medicine (SMFM) to adopt care models that incorporate medical complexity, psychosocial context, and patient preferences [179–181]. Both organizations have issued calls to expand the use of proven interventions, particularly among high-risk and underserved populations [182,183]. Importantly, a focus on actionable risk does not negate the relevance of molecular, genetic, or other biologic pathways for PTB prediction [184–186]; rather, it emphasizes that optimizing the use of known, effective interventions in response to modifiable risk represents an essential and scalable starting point.

In this context, our objective in the present study was to develop and evaluate a Preterm Birth Actionable Risk Index (PTB-ARIx) using data from over 1.9 million births in California between 2016 and 2020. The PTB-ARIx was developed to identify and stratify pregnant individuals based on actionable risk factors. We evaluated model performance across PTB subtypes, including spontaneous, medically-indicated, early (<32 weeks), and late (32–36 weeks) PTB, as well as for PTB co-occurring with other adverse outcomes, such as preeclampsia and/or LBW. We also examined care engagement indicators, including timing of prenatal care initiation and number of visits, as potential risk modifiers. Model performance and potential effect modification were assessed overall and within key sociodemographic subgroups with the goal of informing the development of an equity-oriented clinical tool that improves PTB risk identification and facilitates the uptake of targeted, evidence-based interventions, particularly in those at highest risk.

## Methods

### Study Design and Population

We conducted a retrospective population-based cohort study of singleton live births in California from January 1, 2016, through December 31, 2020. The initial sample included 2,290,649 live births and was restricted to those with gestational ages from 22 to 42 weeks (n = 2,280,142), who were singletons (n = 2,209,857), and had linked birth certificates and pregnant-person/infant hospital discharge records for the birth (n = 1,907,085) (Supplemental Figure 1). Records were derived from California birth certificates maintained by the California Department of Public Health-Vital Records [187], with linkage to pregnant-person and infant hospital discharge records maintained by the California Department of Health Care Access and Information (HCAI) [188]. Linkage was performed using deterministic and probabilistic algorithms (as described in Baer and colleagues [189]) and achieved a linkage rate of 86.3%. The cohort was split into a training sample (births from 2016–2019; n = 1,568,976) and a temporally distinct testing sample (births from 2020; n = 338,109).

### Risk Factors and Predictor Variables

The study examined associations between PTB and 18 actionable clinical, behavioral, and social risk factors (17 single risk factors along with composite risks for preeclampsia), selected for their established links to PTB and the availability of potential evidence-based interventions (Table 1). All variables were derived from birth certificates and/or maternal or infant hospital discharge records, with ICD-10 coding applied where applicable [190,191] (see Supplemental Table 2). Risk factors were modeled as dichotomous variables, except for composite indicators related to elevated preeclampsia risk, which were constructed in alignment with the U.S. Preventive Services Task Force (USPSTF) recommendations for aspirin use in pregnancy [38]. Based on task force recommendations [38], individuals were classified as: 1) having one or more high-risk factors for preeclampsia (pregestational diabetes [type 1 or 2], chronic hypertension, kidney disease, or autoimmune disorders [e.g., systemic lupus erythematosus, rheumatoid arthritis]) only (not having two or more moderate-risk factors for preeclampsia); 2) having two or more moderate-risk factors for preeclampsia (nulliparity, obesity [pre-pregnancy body mass index [BMI] ≥30 kg/m²], advanced maternal age [>34 years], prior adverse pregnancy outcome [e.g., miscarriage, stillbirth, or PTB], interpregnancy interval [IPI] >10 years, conception via in vitro fertilization [IVF], low income [proxied by public insurance enrollment], and/or Black/African American race or ethnicity, and not having one or more high-risk factor for preeclampsia); or 3) having at least one high-risk factor and two or more moderate-risk factors.

**Table 1.**
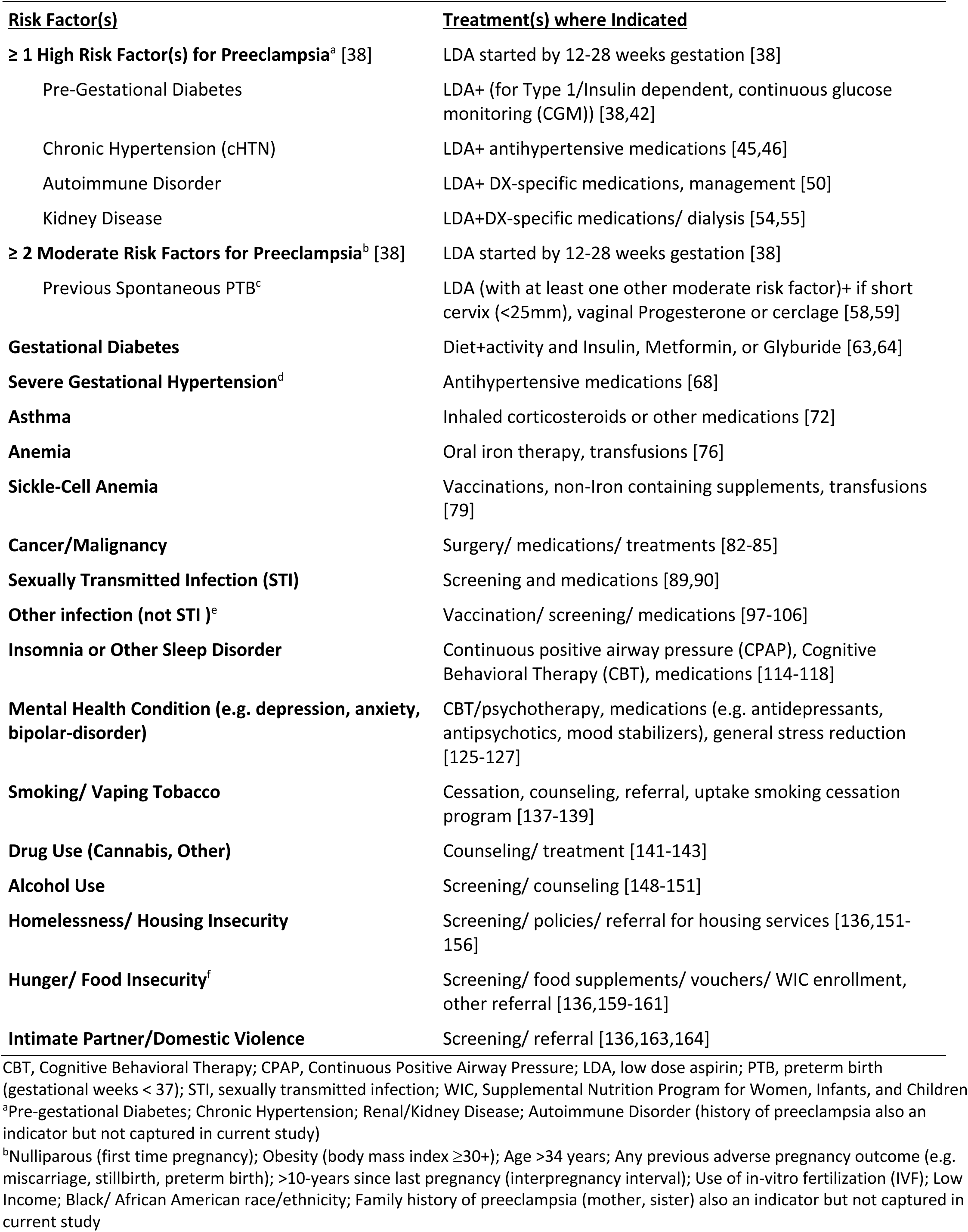
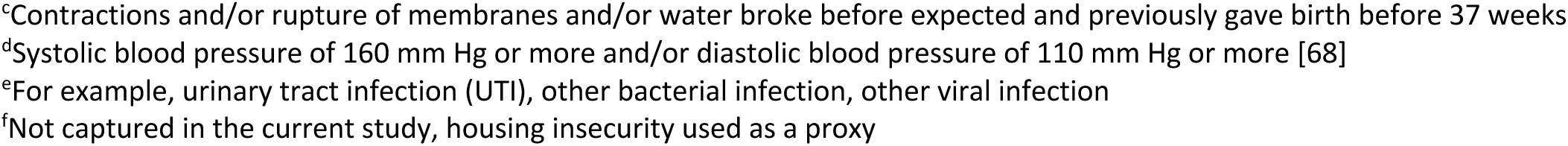
Actionable risk factors for preterm birth considered in the present study and known treatments.

| **<u>Risk Factor(s)</u>** | **<u>Treatment(s) where Indicated</u>** |
| --- | --- |
| **≥ 1 High Risk Factor(s) for Preeclampsia<sup>a</sup> [38]** | LDA started by 12-28 weeks gestation [38] |
| Pre-Gestational Diabetes | LDA+ (for Type 1/Insulin dependent, continuous glucose monitoring (CGM)) [38,42] |
| Chronic Hypertension (cHTN) | LDA+ antihypertensive medications [45,46] |
| Autoimmune Disorder | LDA+ DX-specific medications, management [50] |
| Kidney Disease | LDA+DX-specific medications/ dialysis [54,55] |
| **≥ 2 Moderate Risk Factors for Preeclampsia<sup>b</sup> [38]** | LDA started by 12-28 weeks gestation [38] |
| Previous Spontaneous PTB<sup>c</sup> | LDA (with at least one other moderate risk factor)+ if short cervix (<25mm), vaginal Progesterone or cerclage [58,59] |
| **Gestational Diabetes** | Diet+activity and Insulin, Metformin, or Glyburide [63,64] |
| **Severe Gestational Hypertension<sup>d</sup>** | Antihypertensive medications [68] |
| **Asthma** | Inhaled corticosteroids or other medications [72] |
| **Anemia** | Oral iron therapy, transfusions [76] |
| **Sickle-Cell Anemia** | Vaccinations, non-Iron containing supplements, transfusions [79] |
| **Cancer/Malignancy** | Surgery/ medications/ treatments [82-85] |
| **Sexually Transmitted Infection (STI)** | Screening and medications [89,90] |
| **Other infection (not STI )<sup>e</sup>** | Vaccination/ screening/ medications [97-106] |
| **Insomnia or Other Sleep Disorder** | Continuous positive airway pressure (CPAP), Cognitive Behavioral Therapy (CBT), medications [114-118] |
| **Mental Health Condition (e.g. depression, anxiety, bipolar-disorder)** | CBT/psychotherapy, medications (e.g. antidepressants, antipsychotics, mood stabilizers), general stress reduction [125-127] |
| **Smoking/ Vaping Tobacco** | Cessation, counseling, referral, uptake smoking cessation program [137-139] |
| **Drug Use (Cannabis, Other)** | Counseling/ treatment [141-143] |
| **Alcohol Use** | Screening/ counseling [148-151] |
| **Homelessness/ Housing Insecurity** | Screening/ policies/ referral for housing services [136,151-156] |
| **Hunger/ Food Insecurity<sup>f</sup>** | Screening/ food supplements/ vouchers/ WIC enrollment, other referral [136,159-161] |
| **Intimate Partner/Domestic Violence** | Screening/ referral [136,163,164] |

CBT, Cognitive Behavioral Therapy; CPAP, Continuous Positive Airway Pressure; LDA, low dose aspirin; PTB, preterm birth (gestational weeks < 37); STI, sexually transmitted infection; WIC, Supplemental Nutrition Program for Women, Infants, and Children
<sup>a</sup>Pre-gestational Diabetes; Chronic Hypertension; Renal/Kidney Disease; Autoimmune Disorder (history of preeclampsia also an indicator but not captured in current study)
<sup>b</sup>Nulliparous (first time pregnancy); Obesity (body mass index ≥30+); Age >34 years; Any previous adverse pregnancy outcome (e.g. miscarriage, stillbirth, preterm birth); >10-years since last pregnancy (interpregnancy interval); Use of in-vitro fertilization (IVF); Low Income; Black/ African American race/ethnicity; Family history of preeclampsia (mother, sister) also an indicator but not captured in current study
<sup>c</sup>Contractions and/or rupture of membranes and/or water broke before expected and previously gave birth before 37 weeks
<sup>d</sup>Systolic blood pressure of 160 mm Hg or more and/or diastolic blood pressure of 110 mm Hg or more [68]
<sup>e</sup>For example, urinary tract infection (UTI), other bacterial infection, other viral infection
<sup>f</sup>Not captured in the current study, housing insecurity used as a proxy

These composite variables were further evaluated to examine the relationship between PTB and the number of high-risk and/or moderate-risk factors present modeled as continuous predictors. Of note, prior preeclampsia and family history of preeclampsia (e.g., in a mother or sister), while included in the task force guidelines, are not captured in California birth certificates or hospital discharge records and were therefore excluded from the present analysis. Each of the following risk factors was assessed individually as a binary yes/no variable based on the presence of an ICD-10 diagnosis present in the hospital discharge record [191] (see Supplemental Table for coding): gestational diabetes, gestational hypertension, asthma, anemia (non-sickle cell), sickle cell anemia, malignancy or active cancer diagnosis, sexually transmitted infection (STI, e.g., syphilis, chlamydia, human immunodeficiency virus [HIV]), other infections (e.g., urinary tract infection [UTI], influenza, COVID-19), sleep disorders (e.g., insomnia, obstructive sleep apnea), mental health conditions (e.g., depression, anxiety, bipolar disorder), tobacco use (smoking or vaping), cannabis (marijuana) or other drug use, alcohol use, homelessness or housing insecurity, and exposure to intimate partner violence or domestic violence.

Although there is an ICD-10 code that captures food insecurity [191], it is not consistently used across California hospitals and as such, while it was included in the conceptual framework for the predictive metric given its consistent association with PTB and the demonstrated efficacy of interventions such as food assistance in reducing risk [136,159–161], specific risk-related associations between food insecurity and outcomes are not provided.

### Other Included Variables

Maternal sociodemographic characteristics evaluated included age, race/ethnicity (as self-reported on the birth certificate), education level, and nativity (U.S.-born vs. foreign-born). Race/ethnicity was coded as: Hispanic or NH (where Hispanic = Central or South American; Cuban; Mexican, Mexican American, or Chicano; Puerto Rican; or other Spanish or Hispanic ethnicity), American Indian or Alaska Native, Asian, Black, Native Hawaiian or Other Pacific Islander, White, or other race/ethnicity group (including Asian Indian, Filipino, ≥2 races/ethnicities, other specified race/ethnicity group, refused to state, or unknown race/ethnicity) (see Supplemental Table 2 for additional race/ethnicity coding). Information on prenatal care engagement was also derived from birth certificate records and included trimester of entry into care (first, second, third, or no care), number of prenatal visits (categorized as 0–4, 5–9, or ≥10), and enrollment in the Special Supplemental Nutrition Program for Women, Infants, and Children (WIC) [192].

### Outcome Measures

The primary outcome was PTB, defined as delivery before 37 completed weeks of gestation, based on the best obstetric estimate recorded on the birth certificate. This estimate typically integrates clinical information such as last menstrual period (LMP), early ultrasound findings, and other relevant data [193]. PTBs were further categorized as spontaneous, medically-indicated, or of unknown subtype. Births documented as having “preterm premature rupture of membranes” (PPROM), “preterm labor,” or accompanied by evidence of tocolytic administration were classified as spontaneous PTB. PTBs that were not spontaneous with documentation of “medical induction,” “assisted rupture of membranes,” or cesarean delivery prior to 37 weeks were considered medically-indicated. Individuals with PTB that lacked codes for either spontaneous or medically-indicated categories were classified as unknown subtype (see Supplemental Table 2 for coding).

Predictor-PTB relationships were evaluated in comparison to individuals who had term birth (≥37 weeks) without “other adverse pregnancy outcomes,” defined in this study as birth at ≥37 weeks with one or more of the following: early term birth (37–38 weeks), LBW, small-for-gestational-age birth (SGA; birth weight <10th percentile for gestational age and sex [195]), preeclampsia, “other placental problems” (placenta previa, placental abruption, or placental accreta), SMM [194], major structural congenital anomaly in the infant, maternal death (within 1 year postpartum), or infant death (within 1 year). Additional coding details are provided in Supplemental Table 2.

### Statistical Analysis

Descriptive statistics were generated for both training and testing samples, including distributions of maternal characteristics (age, parity, race/ethnicity, insurance type, education) and rates of PTB. In the training sample, Poisson regression was used to assess the association between each actionable risk factor and PTB. Relative risks (RRs), 95% confidence intervals (CIs), and beta coefficients were reported.

Individual PTB-ARIx scores were computed for use during pregnancy at <20-week and ≥20-week gestation by summing the beta coefficients of risk factors present for each individual in both the training and testing samples. The hospital discharge record did not specify the timing of most risk factors. Therefore, all factors were included in both the <20-week and ≥20-week models, except for diagnoses where timing is specific to the diagnoses (e.g., chronic vs. gestational hypertension, preexisting vs. gestational diabetes). As such, gestational hypertension and gestational diabetes were included only in the ≥20-week model. Additional beta-weighted points were assigned for individuals with more than one high-risk or more than two moderate-risk factors for preeclampsia, based on observed effects from regression models.

The association between per-unit increases in PTB-ARIx score and risk of PTB (vs. term birth without adverse outcome) was evaluated using Poisson regression, with associated RRs and 95% CIs reported. Predictive performance was assessed using receiver operating characteristic (ROC) curves and area under the curve (AUC) metrics. Analyses were stratified by timing of PTB (<32, 32–36 weeks), by PTB subtype (spontaneous vs. medically-indicated), and by co-occurring preeclampsia. PTB-ARIx associations were also examined within race/ethnicity and insurance subgroups. Additional analyses assessed the predictive capacity of PTB-ARIx scores for identifying individuals with other adverse outcomes occurring without PTB.

To examine potential gradients in risk, rates of PTB and other adverse outcomes were compared across PTB-ARIx (<20-week and ≥20-week) score categories: 0.0, >0.0–<1.0, 1.0–<2.0, 2.0–<3.0, and ≥3.0 (based on adding beta coefficients for risk factors present). Poisson regression was used to estimate the risk of PTB or other adverse outcomes compared to term births without complications, with each score group (>0.0) compared to the reference group (score = 0.0).

Although treatment-specific variables were not available, we examined potential mediation by measuring the change in RR in unadjusted versus adjusted Poisson regression models when timing of entry into care (coded 0–3: none, third, second, or first trimester based on level of expected protection) and number of prenatal visits (grouped as <3, 3–6, 7–10, >10; coded 1–4) were included in models examining the relationship between PTB-ARIx score by category (0.0, >0.0–<1.0, 1.0–<2.0, 2.0–<3.0, ≥3.0 vs. 0.0) and PTB. Percentage of potential mediation was calculated as: [RR_unadjusted – RR_adjusted] / [RR_unadjusted – 1] × 100, with 95% CIs obtained using the delta method, based on model-derived variances [196].

All analyses were conducted using SAS version 9.4 (SAS Institute Inc., Cary, NC). The study received ethical approval from the Committee for the Protection of Human Subjects, California Health and Human Services Agency. The study adhered to STROBE (Strengthening the Reporting of Observational Studies in Epidemiology) guidelines [197].

## Results

### Sample Characteristics

In both the training and testing samples, most individuals giving birth were between 18 and 34 years of age (75.9% and 74.2%, respectively), were multiparous (61.5% and 60.6%), and had more than 12 years of education (57.7% and 58.2%). Nearly half identified as Hispanic (47.5% and 47.8%), and over 40% had public insurance (43.5% and 41.5%). The prevalence of PTB was 7.0% in the training sample and 7.2% in the testing sample. More than one-third of individuals in both samples had term deliveries complicated by one or more adverse pregnancy outcomes (34.3% and 35.4%) (Table 2). The most frequent adverse outcomes in those without PTB were early term birth (37–38 weeks; 71.6% and 72.3%) and SGA birth (23.5% and 22.4%) (Supplemental Table 3).

**Table 2.** Sample characteristics, California singleton births in 2016-2019 (training dataset) and in 2020 (testing dataset).

|  | **<u>Training</u>** | **<u>Testing</u>** |
| --- | --- | --- |
|  | n = (%) | n = (%) |
| **Sample** | 1,568,976 (100.0) | 338,109 (100.0) |
| **Maternal Age (Years)** |  |  |
| <18 | 16,283 (1.0) | 2,904 (0.9) |
| 18-34 | 1,191,026 (75.9) | 251,022 (74.2) |
| ≥34 | 361,621 (23.1) | 84,183 (24.9) |
| **Parity** |  |  |
| 0 | 603,985 (38.5) | 133,171 (39.4) |
| ≥1 | 964,991 (61.5) | 204,938 (60.6) |
| **Race/Ethnicity** |  |  |
| Hispanic | 745,823 (47.5) | 161,711 (47.8) |
| Non-Hispanic |  |  |
| American Indian/Alaska Native | 4,751 (0.3) | 1,063 (0.3) |
| Asian | 241,436 (15.4) | 46,618 (13.8) |
| Black/African American | 72,696 (4.6) | 15,775 (4.7) |
| Hawaiian/Pacific Islander | 5,448 (0.4) | 1,118 (0.3) |
| White | 412,411 (26.3) | 89,576 (26.5) |
| Other<sup>a</sup> | 86,411 (5.5) | 22,248 (6.6) |
| ≥2 Race/Ethnicities | 36,244 (2.3) | 7,811 (2.3) |
| **Public Insurance<sup>b</sup>** |  |  |
| Yes | 618,704 (43.5) | 140,288 (41.5) |
| No | 887,272 (56.6) | 197,821 (58.5) |
| **Education (Years)** |  |  |
| <12 | 205,927 (13.1) | 38,247 (11.3) |
| 12 | 376,264 (24.0) | 81,634 (24.1) |
| >12 | 905,930 (57.7) | 196,688 (58.2) |
| **Preterm Birth** |  |  |
| Any (<37 weeks) | 109,526 (7.0) | 24,251 (7.2) |
| <32 weeks | 13,467 (0.9) | 3,177 (0.9) |
| 32-36 weeks | 96,059 (6.1) | 21,074 (6.2) |
| **Term, Adverse Outcome<sup>c</sup>** | 538,651(34.3) | 119,725 (35.4) |
| **Term, No Adverse Outcome** | 920,799 (58.7) | 194,133 (57.4) |

<sup>a</sup>Indian (Asian)', 'Filipino', 'Other-specified', 'Refused to state', 'Unknown'
<sup>b</sup>Health insurance through the state MediCal (California Medicaid) program
<sup>c</sup>Birth at 37 or more weeks, one or more of the following: early term birth (37-38 Weeks), low birth weight (LBW, <2500 grams), small for gestational age (SGA) [195], preeclampsia, other placental problem (previa, abruption, accreta), severe maternal morbidity (SMM) [194], major structural birth defect, maternal death <1 year postpartum, infant death <1 year (additional detail for variable coding including in Supplemental Table 2).

### Candidate Actionable Risk Factors

All candidate actionable risk factors identified for inclusion in the study (Table 1) were associated with an increased risk of PTB in the training sample. Composite preeclampsia-related factors demonstrated the strongest associations. The highest risk observed was in individuals with ≥1 high-risk factor and ≥2 moderate-risk factors for preeclampsia, which occurred in 8.5% of those with PTB compared to 1.4% of those with term birth and no other adverse pregnancy outcome (RR 6.73, 95% CI 6.57, 6.89). The presence of ≥1 high-risk factor without ≥2 moderate-risk factors was also associated with substantial elevation in risk (3.1% vs. 1.0%; RR 4.52, 95% CI 4.35, 4.69). Having ≥2 moderate-risk factors without any high-risk factors conferred a more modest increase in risk (RR 1.91, 95% CI 1.88, 1.94); however, nearly half of PTB cases (49.0% vs. 43.9%) fell into this group. Importantly, each additional high-risk factor for preeclampsia beyond one was associated with nearly a threefold increase in risk (RR 2.88, 95% CI 2.84, 2.92), and each additional moderate-risk factor above two conferred a 35% increase in risk (RR 1.35, 95% CI 1.34, 1.36) (Table 3).

**Table 3.** Association between actionable risk factors and preterm birth: California singleton births 2016-2019 (training)^a^.

|  | **Preterm Birth** | **Term, No Adverse<sup>b</sup>** |  |  |
| --- | --- | --- | --- | --- |
| | n =<br>% | n =<br>% | RR<br>(95% CI) | $\beta$ |
| **<u>Sample</u>** | 109,526<br>(100.0) | 920,799<br>(100.0) |  |  |
| **<u>Risk Factor(s)</u>** |  |  |  |  |
| **$\geq 1$ High Risk Factor(s) for Preeclampsia<sup>c</sup>;**<br>**No Moderate $\geq 2^d$** | 3,358<br>(3.1) | 8,750<br>(1.0) | 4.52<br>(4.35, 4.69) | 1.509 |
| **$\geq 2$ Moderate Risk Factors for**<br>**Preeclampsia<sup>d</sup>; No High Risk<sup>c</sup>** | 53,638<br>(49.0) | 494,331<br>(43.9) | 1.91<br>(1.88, 1.94) | 0.647 |
| **$\geq 1$ High Risk Factor(s) for Preeclampsia<sup>c</sup>;**<br>**$\geq 2$ Moderate Risks<sup>d</sup>** | 9,324<br>(8.5) | 13,259<br>(1.4) | 6.73<br>(6.57, 6.89) | 1.898 |
|  | Mean<br>(SD) | Mean<br>(SD) |  |  |
| **Number of High Risk Factors for**<br>**Preeclampsia<sup>c</sup>** | 0.13<br>(0.40) | 0.03<br>(0.16) | 2.88<br>(2.84, 2.92) | 1.058 |
| **Number of Moderate Risk Factors for $> 2$**<br>**Preeclampsia<sup>d</sup>** | 1.74<br>(0.95) | 1.46<br>(0.89) | 1.35<br>(1.34, 1.36) | 0.300 |
|  | n =<br>% | n =<br>% |  |  |
| **Previous Preterm Birth** | 4,399<br>(4.0) | 5,629<br>(0.6) | 4.26<br>(4.13, 4.39) | 1.449 |
| **Gestational Diabetes** | 14,891<br>(13.6) | 78,826<br>(8.2) | 1.63<br>(1.60, 1.66) | 0.488 |
| **Gestational Hypertension** | 16,907<br>(15.4) | 27,887<br>(3.0) | 4.26<br>(4.13, 4.39) | 1.390 |
| **Asthma** | 8,381<br>(7.7) | 49,387<br>(5.4) | 1.40<br>(1.36, 1.43) | 0.333 |
| **Anemia** | 14,486<br>(13.2) | 94,727<br>(10.3) | 1.29<br>(1.26, 1.31) | 0.251 |
| **Sickle-Cell Anemia** | 74<br>(0.07) | 157<br>(0.02) | 3.02<br>(2.40, 3.79) | 1.104 |
| **Cancer/Malignancy** | 233<br>(0.2) | 451<br>(0.1) | 3.21<br>(2.82, 3.65) | 1.166 |
| **Sexually Transmitted Infection (STI)** | 2,077<br>(1.9) | 13,573<br>(1.5) | 1.25<br>(1.20, 1.31) | 0.226 |
| **Other infection (not STI)<sup>e</sup>** | 18,311<br>(16.7) | 82,444<br>(9.0) | 1.85<br>(1.82, 1.88) | 0.616 |
| **Insomnia or Other Sleep Disorder** | 709<br>(0.7) | 2,096<br>(0.2) | 2.39<br>(2.22, 2.57) | 0.870 |
| **Mental Health Condition (e.g. depression, anxiety)** | 15,958<br>(14.6) | 74,763<br>(8.1) | 1.77<br>(1.74, 1.80) | 0.569 |
| **Smoking/ Vaping Tobacco** | 4,977<br>(4.5) | 19,081<br>(2.1) | 1.99<br>(1.94, 2.05) | 0.689 |
| **Drug Use (Cannabis, Other)** | 6,127<br>(5.6) | 14,616<br>(1.6) | 2.88<br>(2.81, 2.90) | 1.059 |
| **Alcohol Use** | 399<br>(0.4) | 1,368<br>(0.2) | 2.13<br>(1.93, 2.35) | 0.755 |
| **Homelessness/ Housing Insecurity<sup>f</sup>** | 672<br>(0.6) | 1,004<br>(0.1) | 3.79<br>(3.51, 4.09) | 1.352 |
| **Intimate Partner/**<br>**Domestic Violence** | 112<br>(0.8) | 423<br>(0.05) | 1.97<br>(1.64, 2.37) | 0.678 |

β, beta coefficient; CI, confidence interval; RR, relative risk; STI, sexually transmitted infection
<sup>a</sup>Coding schema for summary variables provided in Supplemental Table 2.
<sup>b</sup>Term birth (≥ 37 weeks) without any of the following: early term birth (37–38 weeks), low birthweight (LBW; <2,500 grams), small-for-gestational-age birth (SGA; birth weight <10th percentile for gestational age and sex [195]), preeclampsia, “other placental problems” (placenta previa, placental abruption, or placental accreta), SMM [194], major structural congenital anomaly in the infant, maternal death (<1 year postpartum), or infant death < 1 year (see Supplemental Table 2 for additional coding details).
<sup>c</sup>Pregestational Diabetes; Chronic Hypertension; Renal/Kidney Disease; Autoimmune Disorder (history of preeclampsia also an indicator but not captured in current study)
<sup>d</sup>Nulliparous (first time pregnancy); Obesity (body mass index ≥30+); Age >34 years; Any previous adverse pregnancy outcome (e.g. miscarriage, stillbirth, preterm birth); >10-years since last pregnancy (interpregnancy interval); Use of in-vitro fertilization (IVF); Low Income; Black/ African American race/ethnicity; Family history of preeclampsia (mother, sister) also an indicator but not captured in current study
<sup>e</sup>For example, urinary tract infection (UTI), other bacterial infection, other viral infection
<sup>f</sup>Also used as a proxy for food insecurity

Several additional risk factors were associated with threefold or greater increases in PTB risk. Both prior PTB and gestational hypertension were associated with a fourfold increase in risk (4.0% vs. 0.6%, RR 4.26, 95% CI 4.13, 4.39; and 15.4% vs. 3.0%, RR 4.26, 95% CI 4.13, 4.39, respectively). Although less prevalent, sickle cell anemia, cancer or malignancy, and homelessness or housing insecurity were each associated with more than a threefold increase in risk (Table 3).

While several other risk factors were associated with more moderate elevations in risk (RRs ranging from 1.29 to 1.85, all 95% CIs >1.0), many were relatively common, occurring in more than 10% of premature births. These included gestational diabetes (13.6%), anemia (13.2%), non-STI infections (16.7%), and having one or more mental health conditions (14.6%) (Table 3).

### Cumulative Risk and the PTB-ARIx Score: Patterns and Prediction

More than two-thirds of individuals in the study had at least one actionable risk factor (67.2% and 69.4% in the training and testing samples, respectively; ranges 0–12 and 0–10). Over one-quarter had two or more risk factors (28.1% and 29.4%). The mean number of risk factors present was 1.60 (SD 1.35) for individuals with a PTB, 1.25 (1.18) for individuals with a term birth with another adverse outcome, and 0.97 (0.99) for term births without adverse outcomes in the training sample, and 1.59 (1.37), 1.33 (1.21), and 1.00 (1.00) in the testing sample (Supplemental Table 4).

Each unit increase in the <20-week PTB-ARIx score (calculated as the sum of beta coefficients for each risk factor present, with additional points for >1 high-risk or >2 moderate-risk factors for preeclampsia as described in Methods) was associated with a >60% increased risk of PTB in the training sample (RR 1.67, 95% CI 1.67, 1.68) (Table 4). This association was consistent across insurance-by-race/ethnicity groups, with per-unit risk estimates exceeding 1.60 in nearly all strata. Exceptions included American Indian/Alaska Native, Black, Other race/ethnicity, and ≥2 races/ethnicities with public insurance, where per-unit risks ranged from 1.47 to 1.53 (all CIs >1.0) (Supplemental Table 5).

**Table 4.** Association between per-unit increase in <20– and ≥20-week PTB-ARIx score and risk of preterm birth, preeclampsia, and term birth with other adverse outcomes^a^ (versus term birth without adverse outcome) in the training and testing samples.

| PTB-ARlx Score | <20 Week |  | ≥20 Week |  |
| --- | --- | --- | --- | --- |
|  | Training<br>RR (95% CI) | Testing<br>RR (95% CI) | Training<br>RR (95% CI) | Testing<br>RR (95% CI) |
| **Any (<37 weeks)<sup>a</sup>** | 1.67 (1.67, 1.68) | 1.66 (1.64, 1.68) | 1.72 (1.71, 1.72) | 1.67 (1.66, 1.69) |
| Spontaneous | 1.91 (1.89, 1.93) | 1.99 (1.96, 2.03) | 1.99 (1.97, 2.00) | 2.03 (2.00, 2.07) |
| Medically-Indicated | 2.14 (2.12, 2.16) | 2.27 (2.23, 2.31) | 2.33 (2.32, 2.35) | 2.43 (2.39, 2.47) |
| **<32 weeks<sup>a</sup>** | 2.17 (2.14, 2.19) | 2.08 (2.03, 2.13) | 2.17 (2.15, 2.20) | 2.05 (2.00, 2.10) |
| Spontaneous | 2.36 (2.31, 2.41) | 2.79 (2.72, 2.85) | 2.38 (2.33, 2.42) | 2.43 (2.33, 2.55) |
| Medically-Indicated | 2.00 (1.95, 2.04) | 2.91 (2.77, 3.05) | 3.06 (2.99, 3.12) | 3.11 (2.98, 3.20) |
| **32-36 weeks<sup>a</sup>** | 1.67 (1.66, 1.68) | 1.68 (1.66, 1.70) | 1.73 (1.72, 1.74) | 1.70 (1.68, 1.71) |
| Spontaneous | 1.89 (1.88, 1.91) | 1.99 (1.95, 2.04) | 1.99 (1.97, 2.00) | 2.04 (2.01, 2.08) |
| Medically-Indicated | 2.11 (2.09, 2.13) | 2.27 (2.23, 2.32) | 2.32 (2.30, 2.34) | 2.44 (2.40, 2.42) |
| **Preeclampsia (Any)** | 1.96 (1.95, 1.97) | 1.89 (1.88, 1.92) | 2.44 (2.43, 2.45) | 2.31 (2.29, 2.33) |
| <32 Weeks | 2.91 (2.85, 2.97) | 2.91 (2.79, 3.02) | 3.27 (3.21, 3.32) | 3.27 (3.16, 3.39) |
| 32-36 Weeks | 2.42 (2.40, 2.45) | 2.49 (2.45, 2.54) | 2.82 (2.80, 2.84) | 2.80 (2.76, 2.84) |
| ≥37 Weeks | 1.97 (1.96, 1.98) | 1.90 (1.88, 1.93) | 2.63 (2.62, 2.64) | 2.46 (2.44, 2.48) |
| **Term, Other Adverse<sup>a</sup>** | 1.19 (1.19, 1.20) | 1.21 (1.20, 1.21) | 1.26 (1.26, 1.27) | 1.27 (1.27, 1.28) |

AUC, area under the receiver operating characteristic curve; CI, confidence interval; RR, relative risk; PTB-ARlx, actionable risk index for preterm birth
<sup>a</sup>Includes individuals with spontaneous PTB, medically-indicated PTB, and PTB with an unknown subtype; PTBs with “preterm premature rupture of membranes” (PPROM), “preterm labor,” or accompanied by evidence of tocolytic administration were classified as spontaneous PTB; PTBs without spontaneous PTB but with documentation of “medical induction,” “assisted rupture of membranes,” or cesarean delivery prior to 37 weeks, were considered medically-indicated; PTB without specific codes aligning with either spontaneous or provider-initiated PTB were classified as unknown subtype (see Supplemental Table 2 for specific coding).

Findings with respect to the association between per-unit increase in the <20-week PTB-ARIx score and PTB were similar in the testing sample (RR 1.66, 95% CI 1.64, 1.68) and were also found to be similar with respect to ≥20-week PTB-ARIx score findings. Specifically, each per-unit increase in the ≥20-week PTB-ARIx score was associated with >60% increase in risk for PTB in both the training (RR 1.72, 95% CI 1.71, 1.72) and testing (RR 1.67, 95% CI 1.66, 1.69) samples. Importantly, per-unit increases in both <20– and ≥20-week scores were associated with early (<32 weeks) and late PTB (32–36 weeks), as well as with spontaneous and medically-indicated PTB, and with co-occurring preeclampsia (Table 4). Stronger associations were observed for early PTB, wherein per-unit increases in the <20-week score were associated with more than a twofold increase in PTB risk in both the training and testing samples (RR 2.17, 95% CI 2.14, 2.19; RR 2.08, 95% CI 2.03, 2.13, respectively). The relationship was particularly pronounced for early PTB with preeclampsia, wherein per-unit increases in the <20-week score were associated with nearly a threefold increase in risk (RR 2.91, 95% CI 2.85, 2.97; RR 2.91, 95% CI 2.79, 3.02) and per-unit increases in the ≥20-week score were associated with more than a threefold increase in risk in both the training and testing samples (RR 3.27, 95% CI 3.21, 3.32; RR 3.27, 95% CI 3.16, 3.39).

Performance of the <20– and ≥20-week PTB-ARIx scores by AUC indicated stronger discrimination for PTB <32 weeks. The prediction was particularly robust when PTB <32 weeks co-occurred with preeclampsia. The <20-week PTB-ARIx score showed modest discrimination for late PTB (AUC 0.626, 95% CI 0.625, 0.628, and AUC 0.628, 95% CI 0.623, 0.632 in the training and testing samples, respectively). Performance improved for PTB <32 weeks, with AUCs of 0.691 (95% CI 0.686, 0.696) and 0.676 (95% CI 0.666, 0.686), respectively. Discrimination exceeded 70% for spontaneous and medically-indicated PTB <32 weeks and surpassed 80% when PTB <32 weeks co-occurred with preeclampsia (Supplemental Table 7, Figure 1).

**Figure 1.**
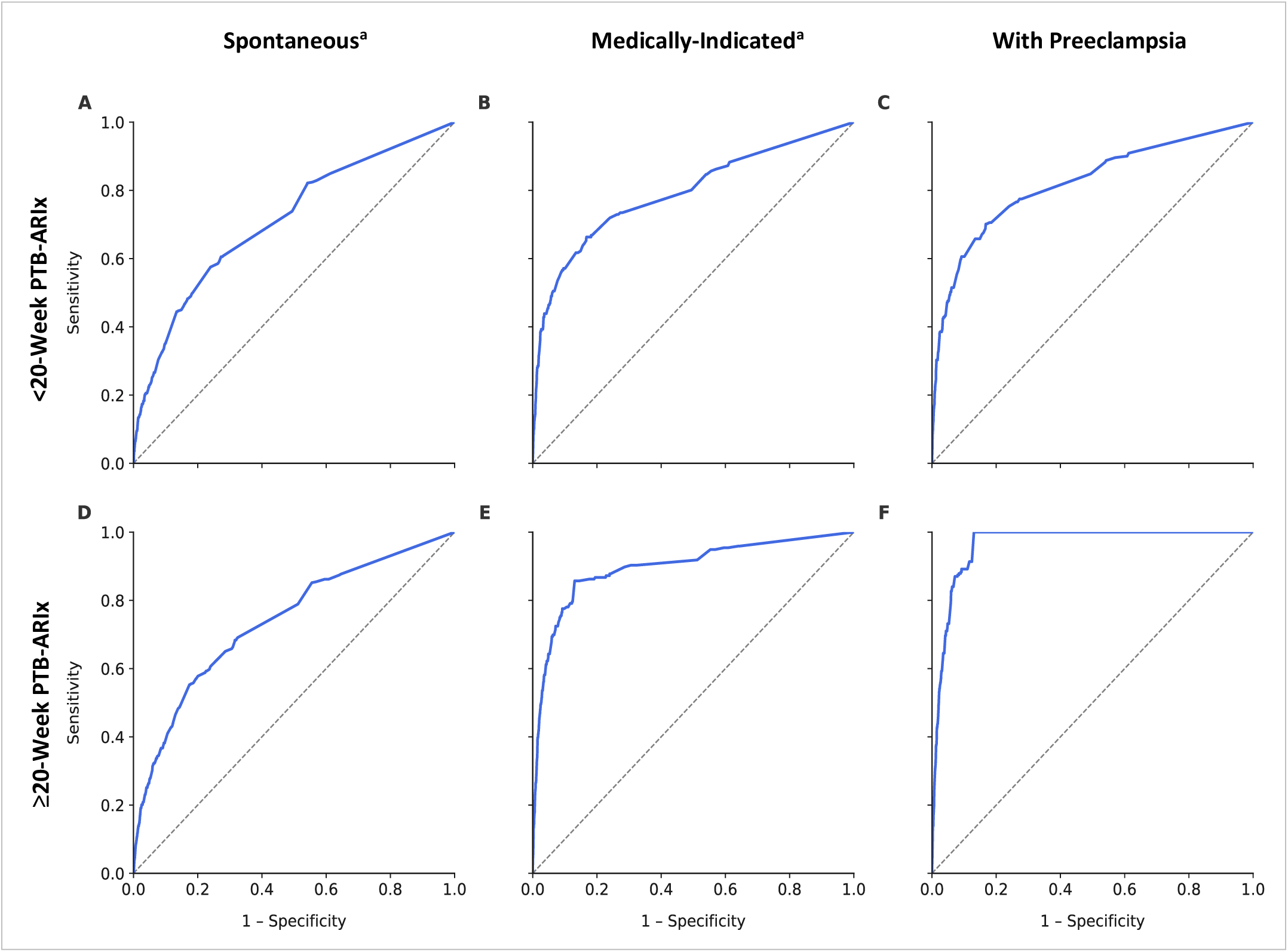
Performance of the <20-week and ≥20-week actionable risk index (PTB-ARIx) in predicting early preterm birth (PTB, <32 weeks) by subtype^a^ and co-occurrence with preeclampsia compared with term births without adverse outcomes^b^ as measured by area under the receiver operating characteristic curve (AUC). **A.** AUC = 0.713, 95% CI 0.704, 0.721; **B.** AUC = 0.774, 95% CI 0.762, 0.785; **C.** AUC = 0.812, 95% CI 0.803, 0.822; **D**. AUC = 0.745, 95% CI 0.737, 0.753; **E.** AUC = 0.880, 95% CI 0.872, 0.889; **F.** AUC = 0.965, 95% CI 0.963, 0.966 AUC, area under the receiver operating characteristic curve; PTB, preterm birth; PTB-ARIx, actionable risk index for preterm birth ^a^PTBs with “preterm premature rupture of membranes” (PPROM), “preterm labor,” or accompanied by evidence of tocolytic administration were classified as spontaneous PTB; PTBs without spontaneous PTB but with documentation of “medical induction,” “assisted rupture of membranes,” or cesarean delivery prior to 37 weeks, were considered medically-indicated (see Supplemental Table 2 for specific coding). ^b^Term birth (≥ 37 weeks) without any of the following: early term birth (37–38 weeks), low birthweight (LBW; <2,500 grams), small-for-gestational-age birth (SGA; birthweight <10th percentile for gestational age and sex [195]), preeclampsia, “other placental problems” (placenta previa, placental abruption, or placental accreta), SMM [194], major structural congenital anomaly in the infant, maternal death (<1 year postpartum), or infant death < 1 year (see Supplemental Table 2 for additional coding details).

The ≥20-week PTB-ARIx score demonstrated stronger performance across outcomes compared to the <20-week score. For spontaneous PTB <32 weeks in the training sample, the AUC was 0.745 (95% CI 0.737, 0.753); for medically-indicated PTB <32 weeks, the AUC was 0.880 (95% CI 0.872, 0.889); and for PTB <32 weeks co-occurring with preeclampsia, the AUC reached 0.965 (95% CI 0.963, 0.966), with similar patterns observed in the testing sample (Supplemental Table 7, Figure 1).

### Risk Groupings and Mediation Analyses

When the associations between the <20– and ≥20-week PTB-ARIx scores, PTB, and term birth accompanied by another adverse pregnancy outcome were examined by risk score groupings (0.01–<1.00, 1.00–<2.00, 2.00–<3.00, ≥3.00) compared with the 0.00 reference group, consistent gradients of increasing risk were observed. For the <20-week PTB-ARIx score, 4.7% of individuals with a score of 0.00 in the training sample experienced PTB, compared with 6.4%, 9.8%, 16.7%, and 26.1% in the 0.01–<1.00, 1.00–<2.00, 2.00–<3.00, and ≥3.00 groups, respectively. RRs rose from 1.41 (95% CI 1.39, 1.43) in the 0.01–<1.00 group to 7.04 (95% CI 6.85, 7.24) in the ≥3.00 group. Similar patterns were observed in the testing sample, with PTB rates ranging from 6.4% to 26.6% among those with scores >0.00 compared with 5.0% in the 0.00 group (RRs 1.32 to 6.99; all 95% CIs >1.00). Associations between <20-week PTB-ARIx groupings and term birth with another adverse outcome were more modest but demonstrated the same pattern. In the training sample, 31.9% of those with a score of 0.00 experienced a term birth with an adverse outcome compared with 46.4% in the ≥3.00 group (RR 1.88, 95% CI 1.84, 1.91), with similar findings seen in the testing sample (Figure 2; Supplemental Table 8).

**Figure 2.**
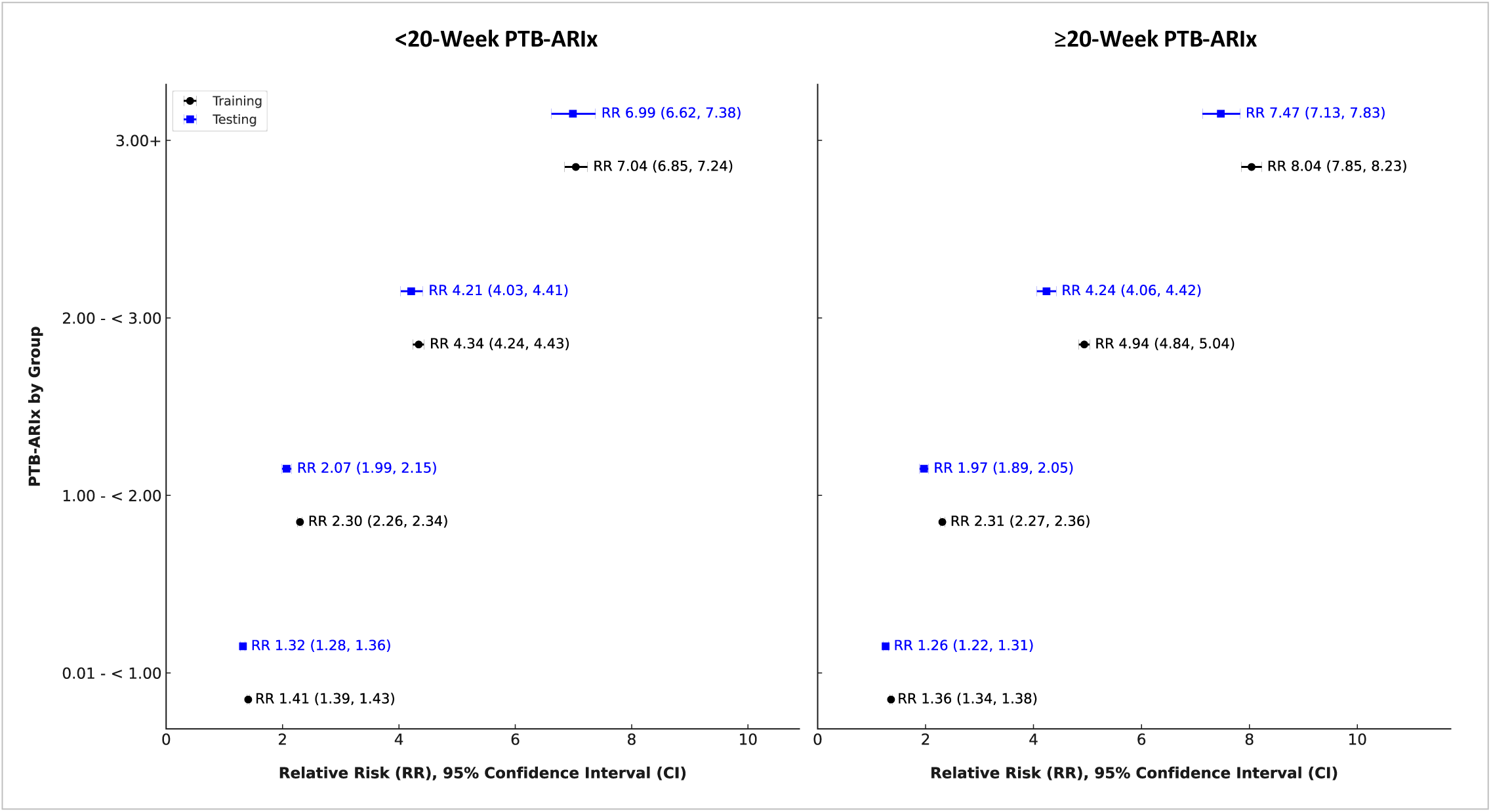
Risk for preterm birth (PTB) by <20– and ≥20-week actionable risk index for PTB (PTB-ARIx) score by group in the training and testing samples. PTB, preterm birth; PTB-ARIx, actionable risk index for preterm birth

In the training sample, 4.3% of individuals with a ≥20-week PTB-ARIx score of 0.00 had a PTB, whereas rates rose to 5.7%, 8.8%, 14.7%, and 23.9% across the 0.01–<1.00, 1.00–<2.00, 2.00–<3.00, and ≥3.00 groups, respectively. Corresponding RRs ranged from 1.36 (95% CI 1.34, 1.38) in the 0.01–<1.00 group to 8.04 (95% CI 7.85, 8.23) in the ≥3.00 group. In the testing sample, PTB rates increased from 4.8% in the 0.00 group to 5.9%–23.2% across higher groupings, with RRs of 1.26 to 7.47 (all 95% CIs >1.00). For term births complicated by another adverse outcome, risk patterns were more modest but still graded. In the training sample, 30.1% of those with a score of 0.00 experienced a term birth with an adverse outcome, compared with 51.5% in the ≥3.00 group (RR 2.15, 95% CI 2.12, 2.18). A similar gradient was evident in the testing sample (Figures 2–3; Supplemental Table 8).

**Figure 3.**
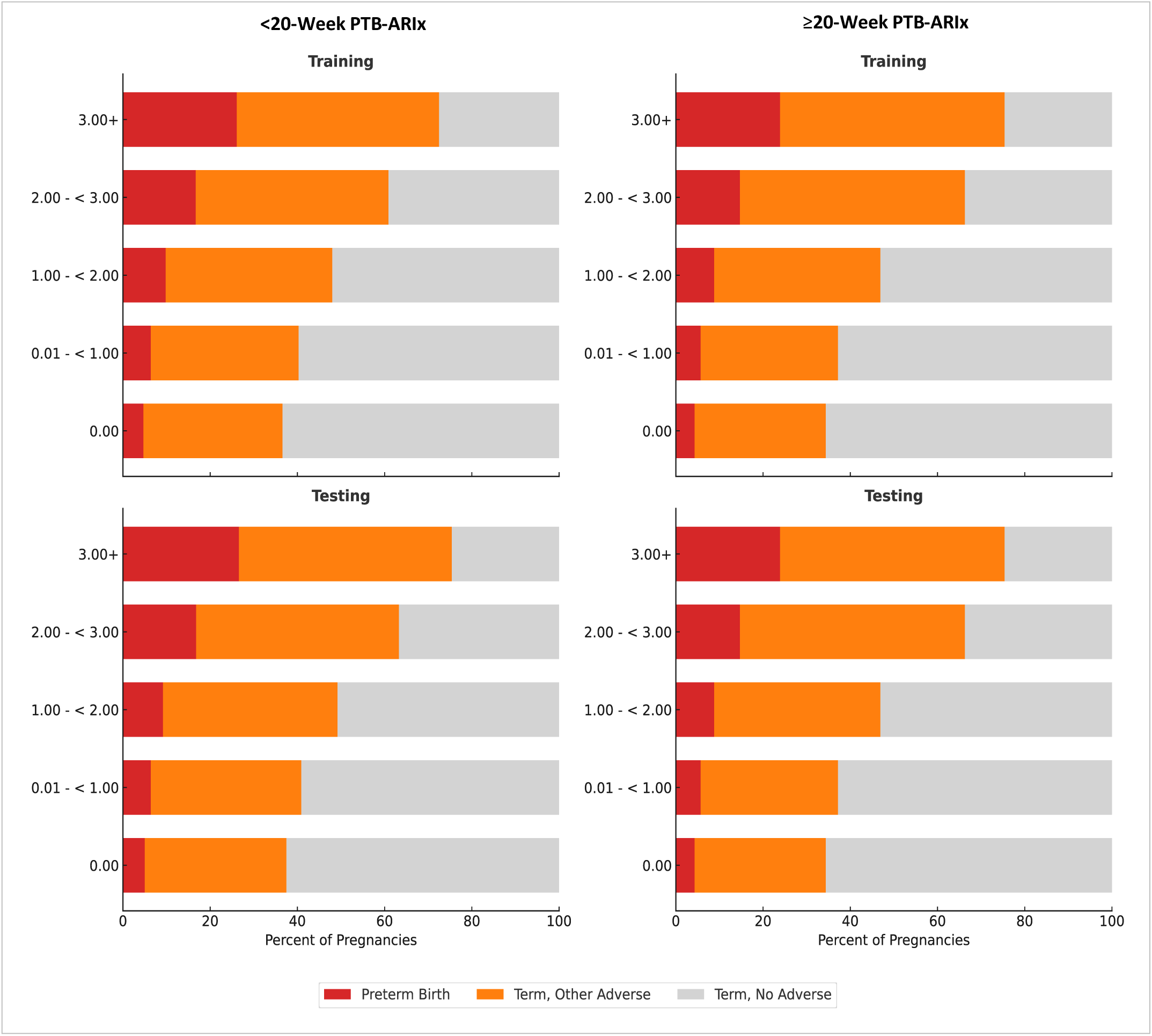
Individuals with preterm birth (PTB), term birth with other adverse outcomes^a^, or term birth without other adverse outcomes by <20– and ≥20-week actionable risk index for PTB (PTB-ARIx) score by group in the training and testing samples. PTB, preterm birth; PTB-ARIx, actionable risk index for preterm birth ^a^Term birth (≥37 weeks) with any of the following: early term birth (37–38 weeks), low birth weight (LBW, <2,500 grams), small-for-gestational-age birth (SGA; birthweight <10th percentile for gestational age and sex [195]), preeclampsia, “other placental problems” (placenta previa, placental abruption, or placental accreta), SMM [194], major structural congenital anomaly in the infant, maternal death (<1 year postpartum), or infant death < 1 year (see Supplemental Table 2 for additional coding details).

Adjusting models for timing of entry into care (coded 0–3, where 0 = no prenatal care and 1–3 = third, second, and first trimesters, respectively) did not change the observed associations between <20– and ≥20-week PTB-ARIx scores and PTB in either training or testing subsets (with percentage possibly mediated values ranging from 0.00 to 4.67%, and all lower bounds of the 95% CIs <0.00). In contrast, including number of prenatal visits (coded 1–4, corresponding to <3, 3–6, 7–10, and 11+ visits) demonstrated robust associations, particularly for the ≥3.00 versus 0.00 PTB-ARIx score comparisons. For example, in the <20-week PTB-ARIx model for PTB in the training sample, the RR decreased from 7.04 (95% CI 6.85, 7.24) to 5.11 (95% CI 4.96, 5.26), corresponding to 31.95% possibly mediated (95% CI 28.64%, 35.27%). In the ≥20-week PTB-ARIx model, the RR decreased from 8.04 (95% CI 7.85, 8.23) to 6.38 (95% CI 6.22, 6.53), with 23.58% possibly mediated (95% CI 20.56%, 26.60%) (Figure 4; Supplemental Table 9). Similar patterns were observed in the testing sample (Figure 4; Supplemental Table 9).

**Figure 4.**
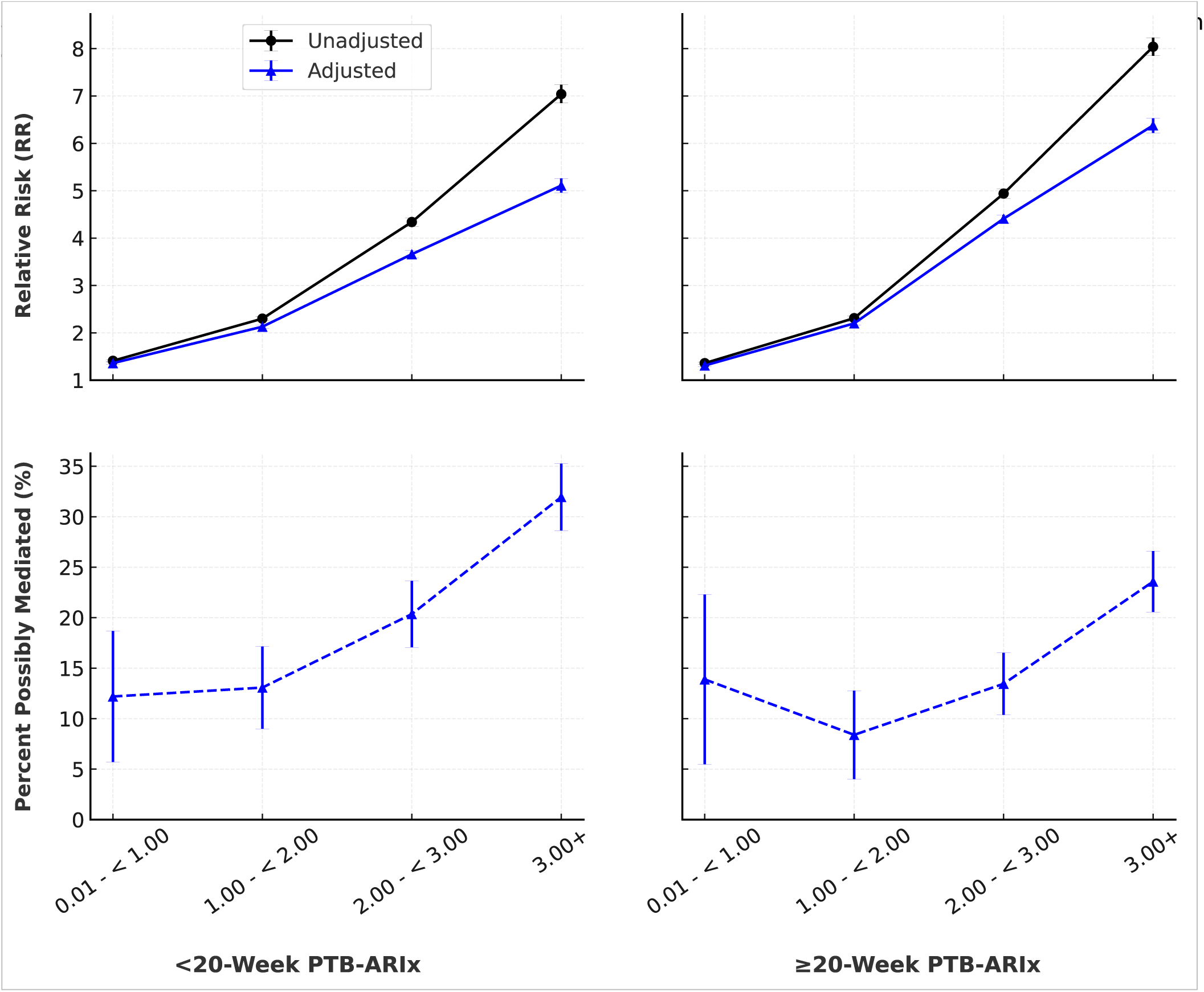
Risk of preterm birth (PTB) versus term birth without adverse pregnancy outcome^a^ by <20– and ≥20-week. PTB, preterm birth; PTB-ARIx, actionable risk index for preterm birth; RR, relative risk ^a^Term birth (≥ 37 weeks) without any of the following: early term birth (37–38 weeks), low birthweight (LBW; <2,500 grams), small-for-gestational-age birth (SGA; birthweight <10th percentile for gestational age and sex [195]), preeclampsia, “other placental problems” (placenta previa, placental abruption, or placental accreta), SMM [194], major structural congenital anomaly in the infant, maternal death (<1 year postpartum), or infant death < 1 year (see Supplemental Table 2 for additional coding details). ^b^Adjusted for number of prenatal visits grouped as <3, 3-6, 7-10, 11+ and coded as 1-4 ^c^Where percent possibly mediated by prenatal visits calculated as (relative risk (RR) (unadjusted) – RR (adjusted)) / (RR (unadjusted − 1)) × 100.

## Discussion

In this large, population-based study, we developed and tested the PTB-ARIx, a tool designed to identify pregnant individuals at increased risk for PTB based on actionable risk factors observed before and after 20 weeks’ gestation. In this context, the PTB-ARIx demonstrated consistent associations with PTB overall and across subtypes, with the strongest predictive power observed for early PTB and for early PTB co-occurring with preeclampsia. Notably, we found that each unit increase in the <20-week and ≥20-week PTB-ARIx scores was associated with more than a 60% elevation in PTB risk, wherein effect sizes were found to increase as gestational age at delivery decreased and when PTB co-occurred with preeclampsia. For early PTB, per-unit increases in the <20-week score conferred more than a twofold increase in risk, and risks approached threefold for early PTB complicated by preeclampsia. Discrimination was modest for late PTB but improved substantially for early PTB, surpassing 0.70 for both spontaneous and medically-indicated subtypes, and exceeding 0.80 when early PTB occurred with preeclampsia. The ≥20-week PTB-ARIx score achieved potent discrimination for early PTB co-occurring with preeclampsia, with AUC values approaching 0.97.

Importantly, when PTB-ARIx scores were stratified by score levels, some groups were found to be at especially increased risk for PTB or for term birth occurring with other adverse outcomes. About three in four of those with a <20-or ≥20-week PTB-ARIx score of ≥3.00 had a PTB or term birth with another adverse outcome in both the training and testing samples. While data were not available on whether individuals received any treatment or intervention related to specific risk factors, when the number of prenatal visits was evaluated as a proxy, findings revealed potentially potent mediation of risk as a function of receipt of care, especially in those with scores at or above 3.00 (with percentage possibly mediated values ranging from about 20% for <20-week PTB-ARIx score and PTB in the testing sample to about 32% for ≥20-week PTB-ARIx score and PTB in the training sample).

### Risk Prediction: PTB-ARIx Compared to Other Metrics

Findings from the present study are consistent with prior investigations in terms of observed associations between specific risk factors and PTB [2,3,34,35,173,177,198]. However, notable differences emerge when comparing this work with other prediction efforts that have relied on clinical, social, and behavioral factors, in terms of factors included in models and in performance, particularly by timing of PTB, subtype, and co-occurrence with preeclampsia [173,175–177,199,200]. Across studies focused on prediction using only clinical and social factors, without additional biomarker testing such as blood pressure, cervical length, or metabolite or cytokine assays, reported AUCs have generally fallen between 0.6 and 0.7 for second-trimester or earlier prediction [173,175–177], with somewhat higher values observed for prediction later in pregnancy [173,176].

Importantly, many prior PTB prediction efforts have relied on nonmodifiable and structurally mediated patient factors (e.g., parity, race/ethnicity, poverty) [173,175–177]. In contrast, our model centers on factors embedded within transparent, existing, actionable interventions prescribed by providers, such as USPSTF recommendations for aspirin prophylaxis [38]. This aligns with priorities identified by both patients and providers for improving risk assessment and communication [167–172]. This framing allows the grouping of certain factors into composites, thereby distinguishing this work from prior models and supporting clearer opportunities for translation into care.

Another key distinction of this study is the evaluation of model performance across PTB subtypes, including early and late PTB, spontaneous and medically-indicated PTB, and PTB co-occurring with preeclampsia, reported in the full study cohort. Prior work has often focused narrowly on spontaneous PTB or PTB with preeclampsia, limiting generalizability. For example, a 2024 review by van Eekhout et al. reported a pooled AUC of 0.61 (95% CI 0.60, 0.64) for first-trimester models of spontaneous PTB using maternal characteristics [201]. By comparison, the <20-week PTB-ARIx score showed stronger performance for spontaneous PTB (AUCs 0.648 and 0.661 in training and testing) and even higher accuracy for spontaneous PTB at <32 weeks (0.712 and 0.717). The same model also demonstrated high predictive value for medically-indicated PTB (AUCs 0.680–0.796) and for PTB with preeclampsia (0.771–0.827), underscoring the importance of evaluating performance across phenotypes.

With respect to the added contribution of biomarkers, prior studies incorporating measures such as cervical length and cytokines (e.g., IL-6 and TNF-α) have consistently reported AUCs exceeding 0.75 for prediction of spontaneous PTB [174]. This pattern of improvement with biomarker integration has also been observed for broader PTB prediction by our group and others [186,202,203]. While the PTB-ARIx is notable for achieving actionable prediction without additional testing, these findings highlight the potential for even greater accuracy when combined with biomarker data, particularly when used to address known gaps in translation to practice. For example, aspirin prophylaxis in women at elevated risk for preeclampsia [204] and the use of psychotropic medications in women with depression [205] have been shown to reduce the levels of inflammation-related biomarkers (e.g., CRP, TNF-α). A deeper understanding of whether and how treatment influences biomarker signaling and PTB risk could yield important insights into mechanisms of intervention impact. Because the PTB-ARIx is based entirely on actionable risk factors, it may serve as a useful framework for comparing biomarker patterns in high-risk individuals who are pregnant with and without treatment.

Of particular importance, the <20-week PTB-ARIx achieved AUCs >0.80 for early PTB with preeclampsia in both training and testing samples, while the ≥20-week PTB-ARIx achieved AUCs >0.95 for both early and late PTB with preeclampsia. These findings align with prior studies demonstrating strong first– and second-trimester prediction of PTB with preeclampsia using clinical and social factors, considered with and without additional biomarkers [206,207]. As mentioned previously, however, the distinguishing feature of the PTB-ARIx is its exclusive focus on actionable risk.

Specifically, for PTB with preeclampsia, the model incorporates composite groups defined by the presence of one or more high-risk factors and/or two or more moderate-risk factors for preeclampsia (consistent with USPSTF recommendations [38]). Within this frame, the PTB-ARIx also accounts for and leverages how risk accumulates when multiple factors are present. To our knowledge, no other model has applied this approach to prediction or risk stratification for PTB with preeclampsia, despite substantial evidence that risk increases when clinical and social factors co-occur [208,209]. Notably, a recent study employed USPSTF guidelines to examine the relationship between high– and moderate-risk factors and the occurrence of preeclampsia with and without aspirin use [210], but investigators did not evaluate patterns of risk when these factors co-occurred, nor did they assess whether or how risk accumulates when risk factors co-occur. Findings from the present study suggest that models seeking to identify individuals at increased risk for preeclampsia would be strengthened by a more robust incorporation of risk co-occurrence and accumulation.

### Risk Stratification and Actionable Inroads for Intervention

Findings from the present study demonstrate that risk for PTB overall, and by timing, subtype, and co-occurrence with preeclampsia increases substantially as <20– and ≥20-week PTB-ARIx scores increase. Given that the indices rely exclusively on actionable risk factors, all score groupings above 0.00 point to the need for action and follow-up. This framing distinguishes the PTB-ARIx from many other PTB prediction efforts that often emphasize cut points or thresholds at which action should be initiated [176,211,212]. By focusing on actionable risk, the PTB-ARIx highlights opportunities to ensure that clinical, social, or behavioral factors linked to elevated risk, and for which evidence-based interventions exist, can be translated into tailored care. This framing redirects focus from a threshold-for-action design to one that ensures that all interventions are implemented equitably and in accordance with patient preferences. Such an approach creates opportunities to strengthen alignment with ACOG and SMFM recommendations to adopt care models that integrate medical complexity, psychosocial context, and patient values [179–181].

Because the PTB-ARIx is anchored in actionable risk, its application extends across all risks and risk groupings, whether an individual has one or multiple factors present. For example, while factors such as anemia, asthma, and STIs were associated with more modest elevations in risk (RRs 1.25–1.40), each is linked to well-established interventions that improve maternal health and have been shown to be associated with a reduction in risk for PTB and/or risk for other adverse pregnancy outcomes when used (e.g., oral iron therapy or transfusion for anemia [76], inhaled corticosteroids for asthma [72], screening and treatment for STIs [89,90]) [74,77,91,92]. Thus, the PTB-ARIx emphasizes not only transparent risk assessment but also the translation of risk into uptake of interventions, creating a pathway to closing care gaps.

Of particular importance in considering the PTB-ARIx is the recognition that **care milieu** profoundly shapes whether risk is communicated, whether interventions are recommended, and ultimately, whether they are adopted. Breakdowns in this feedback loop are starkly exemplified in patterns of recommendation and uptake of aspirin in pregnancy. Despite robust evidence from randomized trials and guideline endorsements, many patients who would benefit from aspirin use are never counseled about it, and many do not initiate use once prescribed [40]. Moreover, pronounced inequities in both prescription and uptake have been documented [41,210]. These patterns underscore the importance of evaluating the PTB-ARIx through a lens that considers not only predictive accuracy but also its potential role in supporting equitable care delivery and reducing disparities in intervention uptake more broadly.

Beyond enhancing knowledge of risk factors and related interventions, the PTB-ARIx has the potential to strengthen understanding of PTB and its early warning signs, thereby supporting sustained engagement with prenatal care and encouraging earlier care-seeking when symptoms arise. Prior work demonstrates that many individuals first learn about treatments such as tocolytics (to delay labor), magnesium sulfate (to protect neurodevelopment), and antenatal corticosteroids (to accelerate fetal lung development) [213] only in emergency settings, contributing to fear, limited preparedness, and challenges in decision-making [214,215]. In contrast, earlier and proactive communication about PTB risk during routine care has been shown to improve preparedness, build trust, and increase acceptance of evidence-based interventions [214–216]. Embedding the PTB-ARIx into prenatal care pathways may therefore not only increase understanding and uptake of interventions linked to specific risk factors and conditions, but also the broader awareness of PTB and its early signs, facilitating timelier initiation of proven therapies that can improve maternal and neonatal outcomes if premature labor occurs.

### Strengths and Limitations

This study has several notable strengths. The use of a large, population-based dataset enabled robust evaluation of PTB risk across subgroups defined by timing, subtype, and co-occurrence with preeclampsia. By centering the PTB-ARIx on actionable risk factors, the study advances a pragmatic framework that directly connects prediction to opportunities for intervention, aligning with professional calls to incorporate medical complexity, psychosocial context, and patient preferences into prenatal care [179–181]. A key methodological strength was our reliance on crude rather than adjusted risks when deriving risk scores. Adjustment of individual factors in risk models can lead to over-adjustment bias, obscuring the true impact of correlated and cumulative exposures on outcomes [217,218]. By using crude risks and their related beta coefficients, the PTB-ARIx better reflects the aggregate contribution of co-occurring actionable factors, thereby capturing the cumulative burden of medical, social, and behavioral risks. This approach reduces the likelihood of underestimating risk associated with multiple intersecting exposures, a limitation inherent in scores based on adjusted associations. Study design also permitted novel analyses of PTB risk in relation to term births complicated by other adverse outcomes. While these findings will require robust follow-up and investigation in and of themselves, they represent findings rarely considered in prior work. Furthermore, the PTB-ARIx demonstrated robustness across insurance types and race/ethnicity groups, supporting the potential for broad generalizability and the opportunity to inform strategies for addressing inequities in adverse pregnancy outcomes.

Several limitations should also be acknowledged. The use of a large administrative database, while enabling population-level analyses, limited the ability to capture detailed clinical and treatment-related information. Measures such as timing of entry into care and number of prenatal visits served only as proxies for the quality and content of care and did not reflect treatment initiation, adherence, or clinical management. This constrains inference about the mechanisms through which actionable risks may be modified. Also critical is that this administrative dataset did not include information about timing of diagnoses and, as such, some factors like infection may have occurred earlier or later in pregnancy and instead were included in both sets of models. Moving forward, it will be critical to examine model performance by timing of exposures. Also of critical importance is the lack of information about the severity of conditions like diabetes and blood pressure. This will be a very important area of investigation moving forward, as what constitutes “actionable risk” in these groups is strongly tied to severity [38,42,45,46,63,64,68].

Although the PTB-ARIx demonstrated consistent performance across diverse subgroups, prospective validation in specific populations and health system settings is needed to confirm generalizability and guide integration into prenatal care. A further limitation is our limited evaluation of individuals with term deliveries complicated by other adverse outcomes (e.g., early term birth, SGA, SMM). In this study, these outcomes were grouped under a single umbrella given their established short– and long-term associations with maternal and infant morbidity [219–227]. However, the relationship between cumulative risk and each outcome remains unclear. While substantial data link individual risk factors (e.g., maternal hypertension, diabetes, SDoH) to these outcomes [228–231], associations with cumulative risk have not been examined in depth. Future work should assess whether the PTB-ARIx can be adapted to improve prediction and uptake of evidence-based interventions related to other target outcomes, or determine whether adaptation of other models or the development of new outcome-specific actionable indexes is warranted [232,233].

### Next Steps

Given persistent inequities in the occurrence of PTB and related risks [3,33–35], building on these findings, next steps should prioritize external validation of the PTB-ARIx across racially/ethnically and sociodemographically diverse populations, with particular attention given to prospective cohorts that allow for more precise evaluation of care processes and quality as well as timing of exposures. Complementary qualitative work with patients and providers will be essential to identify and understand know-do gaps, optimize communication of risk, and ensure interventions are delivered in preference-concordant ways to allow improved uptake by the patients that need them the most.

Evaluation of the PTB-ARIx as a patient-facing tool in clinical or community settings will also be critical to determine its acceptability, feasibility, and potential to improve understanding of risk and uptake of evidence-based interventions. In parallel, integration of PTB-ARIx-defined risk with biomolecular measures offers opportunities both to improve prediction through multimodal models and to uncover mechanistic pathways underlying PTB and related complications.

More broadly, extending the actionable risk framework beyond PTB to other maternal and infant outcomes, including those encompassed in the “term other adverse outcomes” grouping, should be prioritized. Notably, this includes SMM and maternal mortality, where predictive models may similarly benefit from an increased focus on risk factors that are both routinely measured and modifiable [232,233].

### Conclusions

This study introduces the PTB-ARIx as a novel approach for assessing risk for PTB and related adverse outcomes by centering on actionable risk factors, namely those with known evidence-based interventions that can be initiated during pregnancy. By shifting the focus from prediction alone to risk domains where treatment and support may reduce adverse outcomes, the PTB-ARIx highlights an important opportunity to align risk assessment with action. Findings demonstrate robust performance across subgroups and point to the potential for PTB-ARIx to advance more equitable, patient-centered models of prenatal care. Prospective validation will be essential to confirm its clinical utility. At the practice level, the PTB-ARIx may help guide timely identification and intervention, while at the policy level, its emphasis on actionable risk underscores the need to embed medical, structural, social, and behavioral determinants into standard frameworks for maternal health equity and resource allocation.

## Supporting information

Supplemental Content

## Data Availability

All data produced in the present work are contained in the manuscript

