## Supplemental Content for "“Actionable” Risk for Preterm Birth: Patterns and Prediction in California Singleton Births 2016-2020"

Supplemental Figure 1. Sample selection

Supplemental Table 1. Established risk factors for preterm birth with known treatment(s), associated reduction in risk for preterm birth with use (where available), and associated prescription/uptake patterns.

Supplemental Table 2. Coding schema for summary variables (where detail not provided in methods section).

Supplemental Table 3. Frequency of “other adverse outcomes” in individuals giving birth at term in the training and testing samples.

Supplemental Table 4. Percent of individuals with 0 to ≥5 risk factors for preterm birth (PTB)[a] in the training and testing samples.

Supplemental Table 5. Association between per-unit increase in <20- and ≥20-week PTB-ARIx score and risk of preterm birth (versus term birth without adverse outcome) by race/ethnicity with and without public insurance: California births 2016-2020 (n = 1,568,976, training sample).

Supplemental Table 6. Distribution of preterm birth by timing and subtype in the training and testing samples.

Supplemental Table 7. Performance of the <20-week and ≥20-week actionable risk index (PTB-ARIx) in predicting early preterm birth (PTB) by subtype[a] and co-occurrence with preeclampsia compared with term births without adverse outcomes as measured by area under the receiver operating characteristic curve (AUC).

Supplemental Table 8. Association between <20 and ≥20-week actionable risk index (PTB-ARIx) score by group, PTB, term other adverse outcome, and term no adverse outcomes in the training and testing samples.

Supplemental Table 9. Risk of preterm birth (PTB) versus term birth without adverse pregnancy outcome[a] by <20- and ≥20-week actionable risk index for PTB (PTB-ARIx) by group with and without adjustment for timing of entry into care and by number of prenatal visits with calculation of % possibly mediated (PM).

**Supplemental Figure 1.** Sample selection

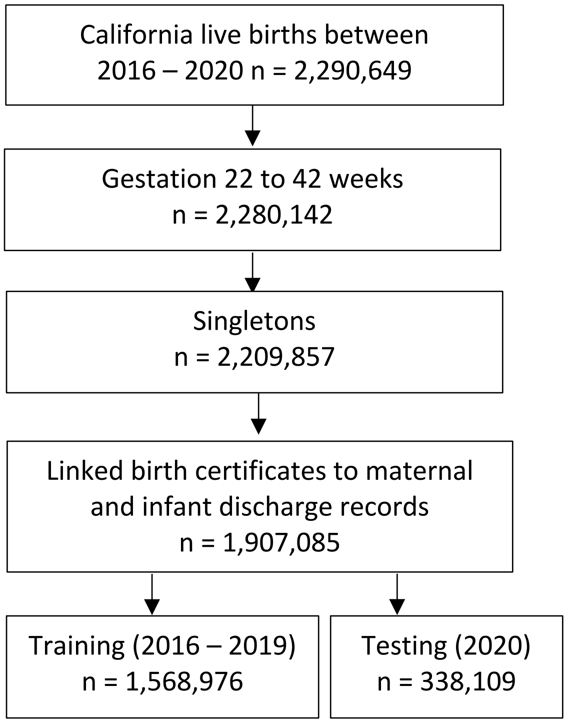

**Supplemental Table 1.** Established risk factors for preterm birth with known treatment(s), associated reduction in risk for preterm birth with use (where available), and associated prescription/uptake patterns.

| Risk Factor(s) | Treatment(s) Where Indicated | Example Studies/ Meta-Analyses: PTB Risk Reduction with Treatment | Example Prescription/ Uptake Patterns |
|---|---|---|---|
| **1 or More High[a] Risk Factor(s) for Preeclampsia** [38] | LDA started by 12-28 weeks gestation [38] | PTB <37 weeks (RR: 0.91, 95% CI 0.87, 0.96); PTB <34 weeks (RR: 0.78, 95% CI 0.61. 0.99) [39] | Prescribed: 1 high risk factor[a]: 57.9% [40]<br><br>Prescribed/ uptake once prescribed:<br>1 high risk factor[a]: 57.7%/ 81.7% [41] |
| **Additional Detail: High Risk Factors for Preeclampsia:** | | | |
| Pre-Gestational Diabetes | LDA+ (for Type 1/Insulin dependent, continuous glucose monitoring (CGM)) [38,42] | LDA: PTB <37 weeks (RR: 0.91, 95% CI 0.87, 0.96); PTB <34 weeks (RR: 0.78, 95% CI 0.61. 0.99) [39] (where pre-gestational diabetes part of a composite indicator)[c]<br><br>CGM: PTB <37 weeks (RR: 0.60, 95% CI 0.39, 0.92) [43] | Prescribed, LDA (any pre-gestational):<br>45.0% [40]<br><br>Prescribed/ uptake once prescribed:<br>53.3%/ 100.0% [41]<br><br>Prescribed, CGM (pre-gestational, type 1): overall 30.0%; Hispanic/ Latina 16.2%; NH White 29.1%; NH Black/ African American 11.3%; Private Insurance 31.1%; Medicaid 9.6% [44] |
| Chronic Hypertension (cHTN) | LDA+ antihypertensive medications [45,46] | LDA: PTB <37 weeks (RR: 0.91, 95% CI 0.87, 0.96); PTB <34 weeks (RR: 0.78, 95% CI 0.61. 0.99) [39] (where cHTN part of a composite indicator)[c]<br><br>LDA (cHTN as the indicator): PTB <37 weeks (OR: 0.63, 95% CI 0.45, 0.89) [47]<br><br>Anti-Hypertensive Medications: Provider-initiated PTB <35 weeks (RR 0.73, 95% CI 0.60, 0.89) [48] | Prescribed, LDA:<br>78.8% [40]<br><br>Prescribed/ uptake of LDA once prescribed:<br>78.8%/ 88.5% [41]<br><br>Prescribed, Antihypertensive Medications:<br>59.4% [49] |
| Autoimmune Disorder | LDA+ DX-specific medications, management [50] | LDA: PTB <37 weeks (RR: 0.91, 95% CI 0.87, 0.96); PTB <34 weeks (RR: 0.78, 95% CI 0.61. 0.99) [39] (where autoimmune disorder part of a composite indicator)[c]<br><br>Hydroxychloroquine (HCQ) for systemic lupus erythematosus | Prescribed, LDA:<br>21.4% [40]<br><br>Prescribed HCQ for SLE:<br>43.0-95.0% [52]<br><br>Prescribed/ uptake of HCQ for SLE once prescribed:<br>49.0%/ 68.6% [53] |

| | | | |
|---|---|---|---|
| | | (SLE): PTB <37 weeks (Pooled OR 0.57, 95% CI 0.46, 0.72) [51] | |
| Kidney Disease | LDA+DX-specific medications/ dialysis, [54,55] | LDA: PTB <37 weeks (RR: 0.91, 95% CI 0.87, 0.96); PTB <34 weeks (RR: 0.78, 95% CI 0.61. 0.99) [39] (where kidney disease part of a composite indicator)[c]<br><br>Dialysis: Within PTB <37 week group, > gestational weeks per hour of dialysis: R2 (meta-regression) = 0.22, 95% CI > 0.0 [56] | Prescribed/ uptake of LDA once prescribed:<br>25.0%/ 100.0% [41]<br><br>Home Hemodialysis versus @Center:<br>All adults, NH Black, NH Asian, Hispanic versus NH White Utilization Ratio (UR) 0.66, 95% CI 0.66, 0.66; 0.77, 95% CI 0.76, 0.77; and 0.65, 95% CI 0.65, 0.65 respectively) [57] |
| **2 or more Moderate[b] Risk Factor(s) for Preeclampsia** [38] | LDA started by 12-28 weeks gestation [38] | PTB <37 weeks (RR: 0.91, 95% CI 0.87, 0.96); PTB <34 weeks (RR: 0.78, 95% CI 0.61. 0.99) [39] | Prescribed: 2 or more moderate risks[b]: 27.3%; Non-Hispanic (NH) Black/ African American+other moderate risk, 32.9% [40]<br><br>Prescribed/ uptake once prescribed:<br>2 or more moderate risks[b]: 4.8%/ 76.9%; NH Black/ African American+other moderate risk: 4.4%/ 50.0%; obese+other moderate risk factor: 5.6%/ 55.6% [41] |
| **Additional Detail: Moderate Risk Factors for Preeclampsia:** | | | |
| Previous Spontaneous PTB[d] | LDA (with at least one other moderate risk factor)+ if short cervix (<25mm), vaginal progesterone or cerclage [58,59] | LDA: PTB <37 weeks (RR: 0.91, 95% CI 0.87, 0.96); PTB <34 weeks (RR: 0.78, 95% CI 0.61. 0.99) [39] (where previous spontaneous PTB occurring with 1 other moderate risk factor for preeclampsia part of a composite indicator)[c]<br><br>Vaginal Progesterone: PTB <35 weeks (RR: 0.68, 95% CI 0.50, 0.93) [60]<br><br>Cerclage: PTB <37 weeks (RR 0.70, 95% CI 0.58, 0.83) [60]<br><br>Vaginal Progesterone+Cerclage: PTB <37 weeks, combined versus progesterone only (RR 0.75, 95% CI 0.58, 0.96), combined versus cerclage only (RR 0.45, 95% CI 0.29, 0.71) [61] | Where previous spontaneous PTB occurring with 1 other moderate risk factor for preeclampsia part of a composite indicator)c<br>LDA Prescribed: Overall 27.3%; NH Black/ African American+other moderate risk, 32.9% [40]<br><br>Where previous PTB (spontaneous not specified, cervix < 25mm) Vaginal Progesterone Treatment: 46.2%; Cerclage (estimate based on exclusion from vaginal progesterone analyses): 6.1% [62] |

| | | | |
|---|---|---|---|
| **Gestational Diabetes** | Diet+activity and Insulin, Metformin, or Glyburide [63,64] | Lifestyle Modification (diet therapy, physical activity), as first-line, escalating to insulin, metformin, or glyburide if glycemic targets not met (mild GDM): PTB <37 weeks considered as part of a composite outcome (RR 0.70, 95% CI 0.60, 0.80) [65] | Insulin, Metformin, or Glyburide Prescribed: 28.0% [66]<br><br>Latina, Medicaid: Insulin, Metformin, or Glyburide Prescribed: 28.8% [67] |
| **Severe Gestational Hypertension**[e] | Antihypertensive medications [68] | Antihypertensive Medications: Preeclampsia including preterm preeclampsia: (RR 0.71, 95% CI 0.54, 0.93) [69] | Antihypertensive Medications Prescribed: 18.6% [70]; Medicaid, 9.9% [71] |
| **Asthma** | Inhaled corticosteroids or other medications [72] | Controlled Versus Poorly Controlled: PTB <37 weeks, (RR 0.76, 95% CI 0.61, 0.94) [73]; (OR 0.85, 95% CI 0.76, 0.96) [74] | Inhaled Corticosteroid/Long-Acting β-Agonist Dispensed for Asthma in the 1st, 2nd, 3rd Trimesters: 59%, 54%, and 52% for privately insured patients; 44%, 38%, and 32% for Medicaid insured [75] |
| **Anemia** | Oral Iron therapy, transfusions [76] | Oral Iron Therapy: PTB <37 weeks (OR 0.59, 95% CI 0.47, 0.72) [77] | Oral or Intravenous Iron Therapy Prescribed/Utilized: 31.1% [78] |
| **Sickle-Cell Anemia** | Vaccinations, non-Iron containing supplements, transfusions [79] | Prophylactic Blood Transfusion(s): PTB <37 weeks, OR 0.67, 95% CI 0.47, 0.97) [80]; Increased Gestational Weeks +2.9 (0.63, 5.17) [81] | Prophylactic Blood Transfusion(s): 29.8% [81] |
| **Cancer/Malignancy** | Surgery/ medications/ treatments [82-85] | Surgery/ Medications/ Treatments , During Versus After Pregnancy: PTB <37 weeks generally found to be higher in those treated during pregnancy versus not but this pattern is lessening over time as treatment is earlier in pregnancy [86] – risk needs to be weighed with consistent findings of reduced risk of maternal death when treatment initiated during versus after pregnancy (RR 0.40, 95% CI 0.20, 0.81) [87] | Surgery/ Medications , During Versus After Pregnancy, Any Chemotherapy or Surgery: 78.0% (12.4% (any chemotherapy), 53.3% (any surgery), 4.2% (both)) [88] |
| **Sexually Transmitted Infection (STI)** | Screening and medications [89,90] | Antibiotics, Active Syphilis: PTB <37 weeks (RR 0.48, 95% CI 0.39, 0.58); Chlamydia: RR 0.58, 95% CI 0.36, 0.93) [91]<br><br>Combination Antiretroviral Therapy (cART), HIV: Spontaneous PTB <37 weeks (HR 0.98, 95% CI 0.96, 0.99) [92] | STI Screening in Keeping with Standards of Care: 76.6% (79.9% NH White, 70.2% Hispanic/Latina, 66.7% NH Black/African American; 80.0% private insurance; 62.7% public/other insurance) [94]<br><br>Screening, Syphilis: 58.0%; Appropriate Use of Antibiotics, Active Syphilis: 12.0% [95] |

| | | | |
|---|---|---|---|
| | | Highly Active Antiretroviral Therapy (HAART), HIV: PTB <28 weeks, <34 weeks, <37 weeks (aRR 0.51, 95% CI 0.31, 0.81; aRR 0.19 , 95% CI 0.08, 0.44; aRR 0.07, 95% CI 0.01, 0.62) [93]<br><br>Monotherapy or Dual Therapy, HIV: PTB <34 weeks (aRR 0.12, 95% CI 0.01, 0.94) [93] | cART, Monotherapy, or Dual Therapy, HIV: 87.7% (86.6% NH Black/African American; 83.5% rural residence) [96] |
| **Other infection (not STI (e.g. urinary tract infection (UTI), other bacterial infection, other viral infection)** | Vaccination/ screening/ medications [97-106] | Vaccination Against COVID-19: PTB <37 weeks (aOR 0.60; 95% CI 0.51, 0.71; [107] aOR 0.74, 95% CI 0.73, 0.75 [108])<br><br>Vaccination Against Influenza: PTB <37 weeks (OR 0.87, 95% CI 0.77, 0.98) [109]<br><br>Screening and Antibiotics for Genital Tract Infection: PTB <37 weeks (RR 0.55, 95% CI 0.41, 0.75) [110]<br><br>Screening and Antibiotics for Asymptomatic UTI: PTB <37 weeks (RR 0.34, 95% CI 0.13, 0.88) [111] | Vaccination Against Influenza: 47.4% (49.9% at or above poverty level; 39.3% below poverty level; 55.5% private/military insurance; 41.4% public insurance; 48.8% White NH; 44.0% Black/African American NH; 48.5% Hispanic; 44.4% aged 18-24; 45.2% aged 25-34; 57.0% aged 35-49) [112]<br>Vaccination Against COVID-19: 30.9% (33.1% at or above poverty level; 23.4% below poverty level; 33.1% private/military insurance; 28.9% public insurance; 27.0% White NH; 29.0% Black/African American NH; 38.6% Hispanic; 32.9% aged 18-24; 28.8% aged 25-34; 34.7% aged 35-49) [112]<br><br>Appropriate Antibiotic Use (Adult Women (pregnancy status unknown)): 45.3% [113] |
| **Insomnia or Other Sleep Disorder** | Continuous positive airway pressure (CPAP), Cognitive Behavioral Therapy (CBT), medications [114-118] | CPAP for obstructive sleep apnea (OSA): Preeclampsia with and without PTB: (RR 0.70, 95% CI 0.50,0.98) [119]<br><br>CBT for Insomnia: Reduced insomnia severity scores difference-in-difference -2.7, 95% CI –4.68, –0.72) [120]<br><br>Digital CBT for Insomnia: Reduced insomnia symptoms time-by-group interaction, difference -0.36; 95% CI -0.48, -0.23 [121]<br><br>Medications for Insomnia/Other Sleep Disorders: Reduced symptoms, some safe in pregnancy and some not [118] | Report of Sleep Problems to Provider (Adults), Insomnia: 40.1%; Snorting, Gasping, Breathing Cessation While Sleeping (symptoms of obstructive sleep apnea): 28.9% [122]<br><br>Adequate digital CBT dose (4 or more sessions) once prescribed for insomnia during pregnancy (RCT): 82.1% (66.6% NH Black/African American; 91.7% NH White) [123]<br><br>CPAP for obstructive sleep apnea in pregnancy, adherence: 54.0% [124] |

| | | | |
|---|---|---|---|
| **Mental Health Condition (e.g. depression, anxiety, bipolar-disorder)** | CBT/psychotherapy, medications (e.g. antidepressants, antipsychotics, mood stabilizers), general stress reduction [125-127] | Psychotherapy for Depression: PTB <37 weeks (HR 0.82, 95% CI 0.71, 0.96) [128]; Each standard deviation of decrease in depressive symptoms (decrease > in therapy group) associated with 3-4 day increase in gestational weeks [129]<br><br>Stress-Reducing Interventions/ Therapy (Pilates, Yoga, Music, Mindfulness, Education): PTB <37 weeks (RR 0.50, 95% CI 0.35, 0.71) [130]<br><br>Digital Therapy (yes versus no by diagnostic grouping): Symptom Reduction, No Known Adverse Impact on PTB or Other Birth Outcomes (measured and no impact or not measured), symptoms of depression (Hedges's g -0.49, 95% CI -0.75, -0.23); symptoms of anxiety (Hedges's g -0.24, 95% CI -0.42, -0.07); symptoms of stress (Hedges's g -0.47, 95% CI -0.79, -0.15) [131]<br>Therapy (yes versus no) by diagnostic grouping: for depression, anxiety, PTSD, Bipolar Disorder, Psychosis and other mental health disorders during pregnancy and during the perinatal period, symptom reduction established, no known adverse impact on PTB or other birth outcomes (measured and no impact or not measured) [132, 133]<br><br>Medications with and without Therapy (yes versus no) by diagnostic grouping: for depression, anxiety, PTSD, Bipolar Disorder, Psychosis and other mental health disorders during pregnancy, symptom reduction established, use based on symptom severity and | Psychotherapy for Depression: 51.0% (63.6% aged 18-24; 50.0% aged 25-34; 43.5% aged 35 years+; 47.2% NH White; 68.6% NH Black; 52.6% NH Hispanic) [128]<br><br>Any Treatment for Mental Health/Substance Use Disorder (U.S. Sample): 33.3% (19.1% NH, Black/African American; 22.2% Hispanic; 40.7% NH White); Insured > twice as likely to receive treatment compared to uninsured (aOR 2.6, 95% CI 1.2, 5.2) [135] |

| | | | |
|---|---|---|---|
| | | medication type [126,128,132-134] | |
| **Smoking/ Vaping Tobacco** | Cessation, counseling, referral, uptake smoking cessation program [137-139] | Smoking Cessation (US): PTB <37 weeks (OR 0.77, 95% CI 0.62, 0.96) [137] | Smoking Cessation: 53.9% (US) (38.3% West Virginia to 79.5% New York City) [137]<br><br>Prolonged Smoking Cessation After Intervention: (8 weeks) 37.0% 95% CI 34.0, 44.0 [140] |
| **Drug Use (Cannabis, Other)** | Counseling/ treatment [141-143] | Treatment Program Participation, Substance Use Disorder: PTB <37 weeks (RR 0.37, 95% CI 0.21, 0.65) [144]<br><br>Cannabis Cessation: PTB <37 weeks (RR 0.37, 95% CI 0.14, 0.97) [145]<br><br>Integrated Treatment for Opioid Use Disorder (into prenatal care): PTB <37 weeks (RR 0.45, 95% CI 0.24, 0.85) [146] | Any Treatment for Mental Health/Substance Use Disorder (U.S. Sample): 33.3% (19.1% NH, Black/African American; 22.2% Hispanic; 40.7% NH White); Insured > twice as likely to receive treatment compared to uninsured (aOR 2.6, 95% CI 1.2, 5.2) [135]<br><br>Psychotherapy; Medication for Opioid Use Disorder: 47.6; 47.4% (Medicaid) [147] |
| **Alcohol Use** | Screening/ counseling [148-151] | Brief Intervention/ Counseling Against Use: PTB <37 weeks (OR 0.67, 95% CI 0.46, 0.98) [149] | Screening/ Counseling (US Sample): 80.0% asked about use (53.5% not graduated from high school; 83.4% graduated from high school; 84.5% some college; 82.2% employed; 77.3% unemployed; health insurance 80.4%; no-health insurance 77.0%) 16.0% of users, counseled to reduce or quit drinking [150] |
| **Homelessness/ Housing Insecurity** | Screening/ policies/ referral for Housing Services [136,151-156] | Housing Insecure, Protection Against Eviction: PTB <37 weeks (RR 0.87, 95% CI 0.86, 0.88) [154]<br><br>Availability of Tenant Right to Counsel: PTB <37 weeks, percent decrease pre-, post-right to counsel -0.91 [155]<br><br>Rent Assistance/Housing Stabilization: PTB <37 weeks composite with low birth weight (<2500 grams), pregnancy loss (RR 0.39, 95% CI 0.21, 0.71) [156] | Hospital/Provider Screening for Housing Instability (all patients, US): 27.8% [157]<br><br>Hospital/Provider Screening for “Social Needs” (including housing instability, pregnant patients, California): 39.4% (systematic screening) [158] |
| **Hunger/ Food Insecurity** | Screening/ food supplements/ vouchers/ WIC enrollment, other referral) [136,159-161] | Food Vouchers: PTB <37 weeks (RR 0.63, 95% CI 0.43, 0.93) [160]<br><br>WIC Enrollment, Public Insurance: PTB <37 weeks (RR | Hospital/Provider Screening for Food Insecurity (all patients, US): 29.6% [157]<br><br>Hospital/Provider Screening for “Social Needs” (including food |

| | | | |
|---|---|---|---|
| | | 0.75, 95% CI 0.72, 0.78 NH Black/African American; RR 0.75, 95% CI 0.74, 0.77 Hispanic; RR 0.85, 95% CI 0.82, 0.77 NH White) [34] | insecurity, pregnant patients, California): 39.4% (systematic screening) [158]<br><br>WIC Enrollment (eligible individuals): 43.7% [162] |
| **Intimate Partner/Domestic Violence** | Screening/ referral [136,163,164] | Increased Help-Seeking/Assistance After Positive Screening: >% discussion with provider; >acceptance of referral [165] | Hospital/Provider Screening for Food Insecurity (all patients, US): 56.4% [157]<br><br>Hospital/Provider Screening for “Social Needs” (including intimate partner violence, pregnant patients, California): 39.4% (systematic screening) [158] |

LDA, low dose aspirin; PTB, preterm birth (gestational weeks < 37); STI, sexually transmitted infection; WIC, Supplemental Nutrition Program for Women, Infants, and Children

[a]Pregestational Diabetes; Chronic Hypertension; Renal/Kidney Disease; Autoimmune Disorder; History of preeclampsia

[b]Nulliparous (first time pregnancy); Obesity (body mass index ≥30+); Family history of preeclampsia (mother, sister); Age >34 years; Any previous adverse pregnancy outcome (e.g. miscarriage, stillbirth, preterm birth); >10-years since last pregnancy (interpregnancy interval); Use of in-vitro fertilization (IVF); Low Income; Black/ African American race/ethnicity

[c]LDA-outcome association for indicator composite (one or more listed in “a” above or 2 or more listed in “b” above

[d]Contractions and/or rupture of membranes and/or water broke before expected and previously gave birth before 37 weeks

[e]Systolic blood pressure of 160 mm Hg or more and/or diastolic blood pressure of 110 mm Hg or more [68]

**Supplemental Table 2.** Coding schema for summary variables (where detail not provided in methods section).[a]

| | Maternal<br>**ICD-10** [191] / **Vital Statistics Code** | Infant<br>**ICD-10** [191] |
|---|---|---|
| **Pre-Gestational Diabetes** | O24.0, O24.1, O24.2, O24.3, E10, E11, E12, E13, E14 | |
| **Chronic Hypertension** | O10 | |
| **Autoimmune Disorder**[f] | M05, M07, M08, M32, M45, M09, L40.5, D68.5, D68.6 | |
| **Kidney Disease** | N18 | |
| **Gestational Diabetes** | O24.4 | P70.0 |
| **Gestational Hypertension** | O13 | P00.0 |
| **Asthma**[f] | J45 | |
| **Anemia (non-sickle cell)**[f] | O99.0 | |
| **Sickle Cell Anemia** | D57.0, D57.1, D57.2 | |
| **Cancer/Malignancy**[f] | Any first letter 'C' | |
| **Sexually Transmitted Infection** | O98.1, O98.2, O98.3 | |
| **COVID-19**[e] | U07.1 | |
| **Other Infection** | O23.0, O23.1, O23.2, O23.3, O23.4, O98.0, O98.4, O98.5, O98.6, O98.7, O98.8, O98.9 | |
| **Insomnia/Sleep Disorder**[f] | F51.0, G47.0, G47.3 | |
| **Mental Health Condition**[f] | F2, F3, F4, F5, F6, O99.3 | |
| **Housing Insecurity**[f] | Z59.0, Z59.1 | |
| **Interpersonal Violence**[f] | T74.01XA, T74.11XA, T74.21XA, T74.31XA, T76.01XA, T76.11XA, T76.21XA, T76.31XA, O9A311, O9A312, O9A313, O9A319, O9A32, O9A33, O9A411, O9A412, O9A413, O9A419, O9A42, O9A43, O9A511, O9A512, O9A513, O9A519, O9A52, O9A53 | |
| **Race/Ethnicity Group** (additional information not in methods section) | | |
| Asian | /'Asian-unspecified', 'Asian-specified', 'Asian-Chinese', 'Asian-Japanese', 'Asian-Korean', 'Asian-Vietnamese', 'Asian-Cambodian', 'Asian-Thai', 'Asian-Lao', 'Asian-Hmong' | |
| Hawaiian/Pacific Islander | /'Hawaiian', 'Guamanian', 'Samoan', 'Other Pacific Islander' | |
| Other | /'Indian (Asian)', 'Filipino', 'Other-specified', 'Refused to state', 'Unknown' | |
| **Preterm Birth (PTB) Subtypes** | | |
| Spontaneous PTB with Premature Rupture of the Membranes (PPROM)[b] | O42 | P01.1 |
| Spontaneous PTB with Intact Membranes[c] | O60 | |
| Medically-Indicated PTB (where no spontaneous PTB)[d] | O82, procedure code 10907ZC, 3E033VJ, 3E0P7GC, 10D00Z0, 10D00Z1, 10D00Z2 | P03.4 |
| **Other Adverse Maternal or Infant Outcome** | | |
| Maternal | | |
| Preeclampsia | O11, O14.0, O14.1, O14.2, O14.9, O15 | |
| Placenta Previa | O44 | P02.0 |
| Placental Abruption | O45 | P02.1 |
| Placental Accreta | O43.2 | |
| Severe Maternal Morbidity (SMM)[f] | | |
| Acute Myocardial Infarction | I21.xx, I22.x | |

| | |
|---|---|
| Aneurysm | I71.xx, I79.0 |
| Acute Renal Failure | N17.x, O90.4 |
| Acute Respiratory Distress Syndrome | J80, J95.1, J95.2, J95.3, J95.82x, J96.0x, J96.2x, J96.9x, R06.03, R09.2 |
| Amniotic Fluid Embolism | O88.112, O88.113, O88.119, O88.12, O88.13 |
| Cardiac Arrest / Ventricular Fibrillation | I46.x, I49.0x |
| Conversion of Cardiac Rhythm | 5A12012, 5A2204Z |
| Disseminated Intravascular Coagulation | D65, D68.8, D68.9, O45.002, O45.003, O45.009, O45.012, O45.013, O45.019, O45.022, O45.023, O45.029, O45.092, O45.093, O45.099, O46.002, O46.003, O46.009, O46.012, O46.013, O46.019, O46.022, O46.023, O46.029, O46.092, O46.093, O46.099, O67.0, O72.3 |
| Eclampsia | O15.X |
| Heart Failure / Arrest During Surgery or Procedure | I97.120, I97.121, I97.130, I97.131, I97.710, I97.711 |
| Puerperal Cerebrovascular Disorders | A81.2, G45.x, G46.x, G93.49, H34.0x, I60.xx, I61.xx, I62.xx, I63.00, I63.01x, I63.1xx, I63.2xx, I63.3xx, I63.4xx, I63.5xx, I63.6, I63.8x, I63.9, I65.xx, I66.xx, I67.xx, I68.xx, O22.50, O22.52, O22.53, I97.810, I97.811, I97.820, I97.821, O87.3 |
| Pulmonary Edema / Acute Heart Failure | I50.1, I50.20, I50.21, I50.23, I50.30, I50.31, I50.33, I50.40, I50.41, I50.43, I50.810, I50.811, I50.813, I50.814, I50.82, I50.83, I50.84, I50.89, I50.9, J81.0 |
| Severe Anesthesia Complications | O29.112-O29.119, O29.122-O29.129, O29.192-O29.199, O29.212-O29.219, O29.292-O29.299, O74.0, O74.1, O74.2, O74.3, O89.0x, O89.1, O89.2, T88.2XXA, T88.3XXA |
| Sepsis | A32.7, A40.x, A41.x, I76, O85, O86.04, R65.20, R65.21, T81.12XA, T81.44XA |
| Shock | O75.1, R57.x, T78.2XXA, T81.10XA, T81.11XA, T81.19XA, T88.6XXA |
| Sickle Cell Disease with Crisis | D57.00, D57.01, D57.02, D57.211, D57.212, D57.219, D57.411, D57.412, D57.419, D57.811, D57.812, D57.819 |
| Air and Thrombotic Embolism | I26.x, O88.012-O88.03, O88.212-O88.23, O88.312-O88.33, O88.812-O88.83, T80.0XXA |
| Hysterectomy | 0UT90ZL, 0UT90ZZ, 0UT97ZL, 0UT97ZZ |
| Temporary Tracheostomy | 0B110F4, 0B113F4, 0B114F4 |
| Ventilation | 5A1935Z, 5A1945Z, 5A1955Z |

PPROM, preterm premature rupture of the membranes; PTB, preterm birth (gestational weeks < 37 completed weeks)
[a]Described for variables where less detail is provided in methods section
*Described for variables where detail not provided in methods section.[a] < 37 completed weeks gestation, indication of premature rupture of the membranes.
[b] <37 completed weeks gestation, indication of tocolytic medication (on birth certificate) or preterm labor (in hospital discharge records), no indication of premature rupture of the membranes.
[c] <37 completed weeks gestation, no indication of preterm labor or PROM, with an indication of induction, augmented labor, artificial rupture of membranes, or cesarean delivery on birth certificate records or in hospital discharge records.
[d]Coded for 2020 births forward.
[e]Coded as present during pregnancy but may also have been present prior to pregnancy.
[f]Based on CDC guidelines for the coding of Severe Maternal Morbidity (SMM) [194].

**Supplemental Table 3.** Frequency of "other adverse outcomes" in individuals giving birth at term in the training and testing samples.[a]

| | **Training, No PTB; Other Adverse Outcome**[a] n = (%) | **Testing, No PTB; Other Adverse Outcome**[a] n = (%) |
|---|---|---|
| **Sample** | 538,651 | 119,725 |
| Early Term (37-38 Weeks) | 385,833 (71.6) | 86,521 (72.3) |
| Low Birth Weight (LBW, <2500 grams) | 28,527 (5.3) | 6,265 (5.2) |
| Small for Gestational Age (SGA)[b] | 126,326 (23.5) | 26,784 (22.4) |
| Preeclampsia | 52,618 (9.8) | 14,901 (12.5) |
| Other Placental Problems[c] | 20,055 (3.7) | 4,403 (3.7) |
| Severe Maternal Morbidity (SMM)[d] | 25,596 (4.8) | 5,841 (4.9) |
| Major Structural Congenital Anomaly | 35,106 (6.5) | 6,879 (5.8) |
| Maternal Death < 1 Year Postpartum | 283 (0.1) | 73 (0.1) |
| Infant Death < 1 Year | 2,052 (0.4) | 382 (0.3) |

LBW, low birth weight; PTB, preterm birth (gestational weeks < 37 completed weeks); SMM, Severe Maternal Morbidity; SGA, Small for Gestational Age

[a]Coding for specific variables described in Supplemental Table 2, individuals may have more than one other adverse outcome.

[b]Birth weight for gestational age and sex at birth <10% [195]

[c]Placental previa, abruption, or accreta

[d]Based on CDC guidelines for the coding of Severe Maternal Morbidity (SMM) [194]

**Supplemental Table 4.** Percent of individuals with 0 to ≥5 risk factors for preterm birth (PTB)[a] in the training and testing samples.

| | Training | | | | Testing | | | |
|---|---|---|---|---|---|---|---|---|
| | All | Preterm Birth | Term, Adverse[a] | Term, No Adverse | All | Preterm Birth | Term, Adverse[b] | Term, No Adverse[a] |
| 0 | 32.8 | 20.1 | 28.8 | 36.7 | 30.7 | 20.3 | 26.3 | 34.6 |
| Any | 67.2 | 79.9 | 71.2 | 63.3 | 69.4 | 79.7 | 73.7 | 65.4 |
| 1 | 39.1 | 35.6 | 37.7 | 40.3 | 40.0 | 36.9 | 38.2 | 41.5 |
| 2 | 17.8 | 23.2 | 20.1 | 15.8 | 18.1 | 21.7 | 20.7 | 16.1 |
| 3 | 6.7 | 12.1 | 8.5 | 5.1 | 7.2 | 11.6 | 9.3 | 5.4 |
| 4 | 2.4 | 5.4 | 3.2 | 1.5 | 2.6 | 5.5 | 3.6 | 1.7 |
| ≥5 | 1.2 | 3.6 | 1.7 | 0.6 | 1.4 | 4.0 | 3.6 | 1.7 |
| Mean (SD) | 1.11 (1.11) | 1.60 (1.35) | 1.25 (1.18) | 0.97 (0.99) | 1.16 (1.12) | 1.59 (1.37) | 1.33 (1.21) | 1.00 (1.00) |
| Median | 1.00 | 1.00 | 1.00 | 1.00 | 1.00 | 1.00 | 1.00 | 1.00 |
| Range | 0-12 | 0-11 | 0-12 | 0-10 | 0-10 | 0-9 | 0-10 | 0-9 |

PTB, preterm birth; SD, standard deviation

[a] ≥1 high risk factor(s) for preeclampsia [38], ≥2 moderate risk factors for preeclampsia [38], previous preterm birth (PTB), gestational diabetes, gestational hypertension, asthma, anemia, sickle-cell anemia, cancer/malignancy, sexually transmitted infection (STI), other infection (not STI), insomnia or other sleep disorder, mental health condition (e.g. depression, anxiety), smoking/ vaping tobacco, drug use (cannabis, other), alcohol use, homelessness/ housing insecurity, intimate partner/domestic violence (based on risk described in Table 3 and variable coding described in Supplemental Table 2).

**Supplemental Table 5.** Association between per-unit increase in <20- and ≥20-week PTB-ARIx score and risk of preterm birth (versus term birth without adverse outcome) by race/ethnicity with and without public insurance[a]: California births 2016-2020 (n = 1,568,976, training sample).

| **PTB-ARIx Score** | **< 20 Week** RR (95% CI) | **≥ 20 Week** RR (95% CI) |
|---|---|---|
| **Sample** | 1.67 (1.67, 1.68) | 1.72 (1.71, 1.72) |
| Public | 1.60 (1.59, 1.61) | 1.64 (1.63, 1.65) |
| Not-Public | 1.76 (1.75, 1.78) | 1.82 (1.81, 1.84) |
| **Hispanic** | 1.68 (1.67, 1.69) | 1.74 (1.73, 1.75) |
| Public | 1.67 (1.66, 1.69) | 1.72 (1.71, 1.74) |
| Not-Public | 1.71 (1.68, 1.73) | 1.79 (1.77, 1.81) |
| **Non-Hispanic** | | |
| American Indian/ Alaska Native | 1.54 (1.45, 1.65) | 1.53 (1.45, 1.63) |
| Public | 1.47 (1.35, 1.59) | 1.44 (1.34, 1.55) |
| Not-Public | 1.70 (1.50, 1.94) | 1.79 (1.60, 2.01) |
| Asian | 1.92 (1.89, 1.95) | 1.97 (1.94, 2.00) |
| Public | 1.83 (1.77, 1.89) | 1.86 (1.81, 1.91) |
| Not-Public | 1.97 (1.93, 2.01) | 2.02 (1.99, 2.06) |
| Black | 1.53 (1.51, 1.56) | 1.57 (1.55, 1.60) |
| Public | 1.50 (1.47, 1.53) | 1.53 (1.50, 1.56) |
| Not-Public | 1.61 (1.57, 1.66) | 1.67 (1.63, 1.72) |
| Hawaiian/Pacific Islander | 1.62 (1.51, 1.74) | 1.63 (1.53, 1.74) |
| Public | 1.61 (1.45, 1.79) | 1.61 (1.47, 1.78) |
| Not-Public | 1.66 (1.50, 1.83) | 1.67 (1.52, 1.82) |
| White | 1.75 (1.73, 1.78) | 1.79 (1.77, 1.82) |
| Public | 1.66 (1.62, 1.70) | 1.68 (1.65, 1.72) |
| Not-Public | 1.88 (1.85, 1.92) | 1.79 (1.77, 1.81) |
| Other[b] | 1.61 (1.58, 1.64) | 1.64 (1.61, 1.67) |
| Public | 1.53 (1.49, 1.57) | 1.54 (1.50, 1.57) |
| Not-Public | 1.70 (1.65, 1.74) | 1.75 (1.71, 1.79) |
| **≥2 Race/Ethnicities** | 1.62 (1.51, 1.66) | 1.63 (1.59, 1.67) |
| Public | 1.53 (1.48, 1.59) | 1.54 (1.49, 1.59) |
| Not-Public | 1.71 (1.64, 1.78) | 1.74 (1.68, 1.80) |

[a]Health insurance through the state MediCal (California Medicaid) program
[b]Indian (Asian)', 'Filipino', 'Other-specified', 'Refused to state', 'Unknown'

**Supplemental Table 6.** Distribution of preterm birth by timing and subtype[a] in the training and testing samples.

| | Training | Testing |
|---|---|---|
| **Preterm Birth** | | |
| **Any (<37 weeks)** | 109,526 (7.0) | 24,251 (7.2) |
| Spontaneous | 36,153 (2.3) | 6,084 |
| Medically-Indicated | 22,272 (1.4) | 4,897 |
| Unknown Subtype | 51,101 (3.3) | 13,270 (3.9) |
| **<32 weeks** | 13,467 (0.9) | 3,177 (0.9) |
| Spontaneous | 4,362 (0.3) | 704 (0.2) |
| Medically-Indicated | 2,270 (0.1) | 534 (0.2) |
| Unknown Subtype | 6,835 (0.4) | 1,939 (0.6) |
| **32-36 weeks** | 96,059 (6.1) | 21,074 (6.2) |
| Spontaneous | 31,791 (2.0) | 5,380 (1.6) |
| Medically-Indicated | 20,002 (1.3) | 4,363 (1.3) |
| Unknown Subtype | 44,266 (2.8) | 11,331 (3.4) |

[a]PTBs with "preterm premature rupture of membranes" (PPROM), "preterm labor," or accompanied by evidence of tocolytic administration were classified as spontaneous PTB; PTBs without spontaneous PTB but with documentation of "medical induction," "assisted rupture of membranes," or cesarean delivery prior to 37 weeks, were considered medically-indicated; PTB without specific codes aligning with either spontaneous or provider-initiated PTB were classified as unknown subtype (see Supplemental Table 2 for specific coding).

**Supplemental Table 7.** Performance of the <20-week and ≥20-week actionable risk index (PTB-ARIx) in predicting early preterm birth (PTB) by subtype[a] and co-occurrence with preeclampsia compared with term births without adverse outcomes[a] as measured by area under the receiver operating characteristic curve (AUC).

| | <20-Week PTB-ARIx Score | | ≥20-Week PTB-ARIx Score | |
|---|---|---|---|---|
| | Training AUC (95% CI) | Testing AUC (95% CI) | Training AUC (95% CI) | Testing AUC (95% CI) |
| **Any (<37 weeks)** | 0.634 (0.633, 0.636) | 0.634 (0.630, 0.638) | 0.670 (0.668, 0.672) | 0.662 (0.658, 0.666) |
| Spontaneous | 0.648 (0.645, 0.651) | 0.661 (0.653, 0.669) | 0.691 (0.688, 0.694) | 0.705 (0.697, 0.712) |
| Medically-Indicated | 0.690 (0.686, 0.694) | 0.720 (0.711, 0.728) | 0.781 (0.777, 0.784) | 0.802 (0.794, 0.810) |
| **<32 weeks** | 0.691 (0.686, 0.696) | 0.676 (0.666, 0.686) | 0.719 (0.714, 0.723) | 0.692 (0.682, 0.702) |
| Spontaneous | 0.713 (0.704, 0.721) | 0.717 (0.695, 0.738) | 0.745 (0.737, 0.753) | 0.750 (0.729, 0.771) |
| Medically-Indicated | 0.774 (0.762, 0.785) | 0.796 (0.774, 0.819) | 0.880 (0.872, 0.889) | 0.882 (0.864, 0.900) |
| **32-36 weeks** | 0.626 (0.625, 0.628) | 0.628 (0.623, 0.632) | 0.663 (0.661, 0.665) | 0.658 (0.654, 0.662) |
| Spontaneous | 0.639 (0.636, 0.642) | 0.653 (0.645, 0.662) | 0.684 (0.680, 0.687) | 0.699 (0.691, 0.707) |
| Medically-Indicated | 0.680 (0.676, 0.684) | 0.710 (0.702, 0.719) | 0.770 (0.766, 0.774) | 0.792 (0.784, 0.801) |
| **Preeclampsia (Any)** | 0.715 (0.713, 0.717) | 0.717 (0.713, 0.721) | 0.955 (0.954, 0.955) | 0.955 (0.954, 0.955) |
| <32 Weeks | 0.812 (0.803, 0.822) | 0.827 (0.808, 0.847) | 0.965 (0.963, 0.966) | 0.965 (0.962, 0.968) |
| 32-36 Weeks | 0.771 (0.767, 0.775) | 0.799 (0.791, 0.807) | 0.962 (0.961, 0.962) | 0.961 (0.960, 0.962) |
| ≥37 Weeks | 0.694 (0.692, 0.696) | 0.691 (0.687, 0.696) | 0.952 (0.952, 0.953) | 0.948 (0.947, 0.949) |
| **Term, Other Adverse[a]** | 0.549 (0.548, 0.550) | 0.556 (0.554, 0.696) | 0.583 (0.582, 0.584) | 0.597 (0.595, 0.599) |

AUC, area under the receiver operating characteristic curve; CI, confidence interval; RR, relative risk; PTB-ARIx, actionable risk index for preterm birth

[a]Term birth (≥ 37 weeks) with one or more of the following: early term birth (37–38 weeks), low birthweight (LBW; <2,500 grams), small-for-gestational-age birth (SGA; birthweight <10th percentile for gestational age and sex [195]), preeclampsia, "other placental problems" (placenta previa, placental abruption, or placental accreta), SMM [194], major structural congenital anomaly in the infant, maternal death (<1 year postpartum), or infant death < 1 year (see Supplemental Table 2 for additional coding details).

**Supplemental Table 8.** Association between <20 and ≥20-week actionable risk index (PTB-ARIx) score by group, PTB, term other adverse outcome[a], and term no adverse outcomes in the training and testing samples.

| | Preterm Birth<br>n (%)<br>RR (95% CI) | Term, Other Adverse[a]<br>n (%)<br>RR (95% CI) | Term, No Adverse<br>n (%)<br>RR (95% CI) |
|---|---|---|---|
| **Training** | | | |
| <20-Week PTB-ARIx | | | |
| 0.00 (n = 579,144) | 27,228 (4.7) | 184,555 (31.9) | 367,361 (63.4)<br>Reference |
| 0.01 - <1.00 (n = 693,944) | 44,693 (6.4)<br>1.41 (1.39, 1.43) | 235,017 (33.9)<br>1.08 (1.08, 1.09) | 414,234 (59.7) |
| 1.00 - <2.00 (n = 204,382) | 20,019 (9.8)<br>2.30 (2.26, 2.34) | 78,097 (38.2)<br>1.27 (1.26, 1.28) | 106,266 (52.0) |
| 2.00 - <3.00 (n = 66,903) | 11,173 (16.7)<br>4.34 (4.24, 4.43) | 29,575 (44.2)<br>1.59 (1.57, 1.61) | 26,155 (39.1) |
| 3.00+ (n = 24,603) | 6,413 (26.1)<br>7.04 (6.85, 7.24) | 11,407 (46.4)<br>1.88 (1.84, 1.91) | 6,783 (27.6) |
| ≥20-Week PTB-ARIx | | | |
| 0.00 (n = 514,750) | 22,064 (4.3) | 155,157 (30.1) | 337,529 (65.6)<br>Reference |
| 0.01 - <1.00 (n = 624,781) | 35,654 (5.7)<br>1.36 (1.34, 1.38) | 196,917 (31.5)<br>1.06 (1.05, 1.07) | 392,210 (62.8) |
| 1.00 - <2.00 (n = 259,130) | 22,739 (8.8)<br>2.31 (2.27, 2.36) | 98,787 (38.1)<br>1.33 (1.32, 1.34) | 137,604 (53.1) |
| 2.00 - <3.00 (n = 126,286) | 18,530 (14.7)<br>4.94 (4.84, 5.04) | 65,133 (51.6)<br>1.92 (1.90, 1.94) | 42,623 (33.8) |
| 3.00+ (n = 44,029) | 10,539 (23.9)<br>8.04 (7.85, 8.23) | 22,653 (51.5)<br>2.15 (2.12, 2.18) | 10,833 (24.6) |
| **Testing** | | | |
| <20-Week PTB-ARIx | | | |
| 0.00 (n = 117,478) | 5,897 (5.0) | 38,198 (32.5) | 73,383 (62.5)<br>Reference |
| 0.01 - <1.00 (n = 153,000) | 9,809 (6.4)<br>1.32 (1.28, 1.36) | 52,851 (34.5)<br>1.08 (1.06, 1.09) | 90,340 (59.1) |
| 1.00 - <2.00 (n = 45,123) | 4,157 (9.2)<br>2.07 (1.99, 2.15) | 18,061 (40.0)<br>1.29 (1.27, 1.31) | 22,905 (50.8) |
| 2.00 - <3.00 (n = 16,265) | 2,725 (16.8)<br>4.21 (4.03, 4.41) | 7,570 (46.5)<br>1.63 (1.59, 1.67) | 5,970 (36.7) |
| 3.00+ (n = 6,243) | 1,663 (26.6)<br>6.99 (6.62, 7.38) | 3,045 (48.8)<br>1.94 (1.87, 2.02) | 1,535 (24.6) |
| ≥20-Week PTB-ARIx | | | |

| | | | |
|---|---|---|---|
| 0.00 (n = 103,630) | 4,922 (4.8) | 31,466 (30.4) | 67,242 (64.9) |
| 0.01 - <1.00 (n = 135,491) | 7,970 (5.9)<br>1.26 (1.22, 1.31) | 42,762 (31.6)<br>1.05 (1.04, 1.07) | 84,759 (62.6) |
| 1.00 - <2.00 (n = 56,580) | 4,582 (8.1)<br>1.97 (1.89, 2.05) | 22,517 (39.8)<br>1.36 (1.34, 1.38) | 29,481 (52.1) |
| 2.00 - <3.00 (n = 30,819) | 4,087 (13.3)<br>4.24 (4.06, 4.42) | 16,671 (54.1)<br>1.94 (1.91, 1.98) | 10,061 (32.7) |
| 3.00+ (n = 11,589) | 2,690 (23.2)<br>7.47 (7.13, 7.83) | 6,309 (54.4)<br>2.22 (2.17, 2.29) | 2,590 (22.4) |

CI, confidence interval; PTB, preterm birth; PTB-ARIx, actionable risk index for preterm birth; RR, relative risk

[a]Term birth (≥37 weeks) without any of the following: early term birth (37–38 weeks), low birthweight (LBW; <2,500 grams), small-for-gestational-age birth (SGA; birthweight <10th percentile for gestational age and sex [195]), preeclampsia, "other placental problems" (placenta previa, placental abruption, or placental accreta), SMM [194], major structural congenital anomaly in the infant, maternal death (<1 year postpartum), or infant death < 1 year (see Supplemental Table 2 for additional coding details).

**Supplemental Table 9.** Risk of preterm birth (PTB) versus term birth without adverse pregnancy outcome[a] by <20- and ≥20-week actionable risk index for PTB (PTB-ARIx) by group with and without adjustment for timing of entry into care[b] and by number of prenatal visits[c] with calculation of % possibly mediated (PM)[d].

| | | Entry Into Care[b] | | Number of Prenatal Visits[c] | |
|---|---|---|---|---|---|
| | RR (95% CI) | RR-adj (95% CI) | % PM (95% CI) | RR-adj (95% CI) | % PM (95% CI) |
| **Training** | | | | | |
| <20-Week PTB-ARIx | | | | | |
| 0.01 - <1.00 | 1.41 (1.39, 1.43) | 1.41 (1.39, 1.43) | 0.00 (-6.85, 6.85) | 1.36 (1.34, 1.38) | 12.20 (5.70, 18.69) |
| 1.00 - <2.00 | 2.30 (2.26, 2.34) | 2.28 (2.24, 2.32) | 1.54 (-2.74, 5.82) | 2.13 (2.09, 2.17) | 13.08 (9.00, 17.15) |
| 2.00 - <3.00 | 4.34 (4.24, 4.43) | 4.26 (4.17, 4.36) | 2.40 (-1.55, 6.34) | 3.66 (3.58, 3.74) | 20.36 ( 7.06, 23.66) |
| 3.00+ | 7.04 (6.85, 7.24) | 6.80 (6.62, 7.00) | 3.97 (-0.55, 8.50) | 5.11 (4.96, 5.26) | 31.95 (28.64, 35.27) |
| ≥20-Week PTB-ARIx | | | | | |
| 0.01 - <1.00 | 1.36 (1.34, 1.38) | 1.35 (1.33, 1.38) | 2.78 (-7.05, 12.61) | 1.31 (1.29, 1.34) | 13.89 (5.48, 22.30) |
| 1.00 - <2.00 | 2.31 (2.27, 2.36) | 2.30 (2.26, 2.34) | 0.76 (-4.05, 5.58) | 2.20 (2.16, 2.24) | 8.40 (4.02, 12.78) |
| 2.00 - <3.00 | 4.94 (4.84, 5.04) | 4.89 (4.79, 4.99) | 1.27 (-2.26, 4.80) | 4.41 (4.32, 4.49) | 13.45 (10.37, 16.53) |
| 3.00+ | 8.04 (7.85, 8.23) | 7.83 (7.64, 8.01) | 2.98 (-0.64, 6.60) | 6.38 (6.22, 6.53) | 23.58 (20.56, 26.60) |
| **Testing** | | | | | |
| <20-Week PTB-ARIx | | | | | |
| 0.01 - <1.00 | 1.32 (1.28, 1.36) | 1.31 (1.27, 1.35) | 3.12 (-14.02, 20.27) | 1.27 (1.23, 1.32) | 15.62 (-14.29, 36.11) |
| 1.00 - <2.00 | 2.07 (1.99, 2.15) | 2.04 (1.96, 2.12) | 2.80 (-7.42, 13.03) | 2.04 (1.96, 2.12) | 2.80 (-13.13, 16.52) |
| 2.00 - <3.00 | 4.21 (4.03, 4.41) | 4.12 (3.94, 4.32) | 2.80 (-5.68, 11.29) | 3.67 (3.50, 3.84) | 16.82 (6.27, 26.69) |
| 3.00+ | 6.99 (6.62, 7.38) | 6.71 (6.34, 7.09) | 4.67 (-3.96, 13.31) | 5.42 (5.11, 5.73) | 26.21 (15.84, 35.58) |
| ≥20-Week PTB-ARIx | | | | | |
| 0.01 - <1.00 | 1.26 (1.22, 1.31) | 1.25 (1.21, 1.30) | 3.85 (-22.32, 30.01) | 1.22 (1.18, 1.27) | 15.38 (-22.73, 41.94) |
| 1.00 - <2.00 | 1.97 (1.89, 2.05) | 1.96 (1.88, 2.04) | 1.03 (-10.34, 12.40) | 1.88 (1.81, 1.96) | 9.28 (-7.87, 22.86) |
| 2.00 - <3.00 | 4.24 (4.06, 4.42) | 4.17 (4.00, 4.35) | 2.16 (-5.45, 9.77) | 3.83 (3.67, 3.99) | 12.65 (2.29, 21.93) |
| 3.00+ | 7.47 (7.13, 7.83) | 7.24 (6.90, 7.59) | 3.56 (-3.89, 11.00) | 6.18 (5.89, 6.49) | 19.94 (10.44, 28.40) |

CI, confidence interval; PM, possibly mediated; PTB, preterm birth; PTB-ARIx, actionable risk index for preterm birth; RR, relative risk

[a]Term birth (≥ 37 weeks) without any of the following: early term birth (37–38 weeks), low birthweight (LBW; <2,500 grams), small-for-gestational-age birth (SGA; birthweight <10th percentile for gestational age and sex [195]), preeclampsia, "other placental problems" (placenta previa, placental abruption, or placental accreta), SMM [194], major structural congenital anomaly in the infant, maternal death (<1 year postpartum), or infant death < 1 year (see Supplemental Table 2 for additional coding details).

[b]Adjusted for timing of entry into care none, third trimester, second trimester, first trimester coded as 0-3 based on expected protection

[c]Adjusted for number of prenatal visits grouped as <3, 3-6, 7-10, 11+ and coded as 1-4 based on expected protection

[d]Where percent possibly mediated by prenatal visits calculated as (relative risk (RR) (unadjusted) – RR (adjusted)) / (RR (unadjusted − 1)) × 10
